# IL-21, inflammatory cytokines and hyper polarized CD8^+^ T cells are central players in Lupus immune pathology

**DOI:** 10.1101/2022.10.18.22281177

**Authors:** Soumya Sengupta, Gargee Bhattacharya, Subhasmita Mohanty, Shubham K Shaw, Gajendra M Jogdand, Rohila Jha, Prakash K Barik, Jyoti R Parida, Satish Devadas

**Affiliations:** Institute of Life Sciences, Bhubaneswar, Odisha, India; Regional Centre for Biotechnology (RCB), Faridabad-Gurgaon, Haryana, India; Odisha Arthritis & Rheumatology Centre (OARC), Bhubaneswar, Odisha, India

**Author notes:** Address of Corresponding authors: Dr. Jyoti R Parida, Senior Consultant, Rheumatologist, Odisha Arthritis & Rheumatology Centre (OARC), Saheed Nagar, Bhubaneswar, Odisha, Dr. Satish Devadas (SD), Infectious Disease Biology, Institute of Life Sciences, (Autonomous Institute of Dept of Biotechnology, Govt. of India), Nalco Square, Bhubaneswar Odisha, India.

**Keywords:** SLE, CD8^+^ T Cells, inflammatory cytokines, Reactive Oxygen Species, Antioxidant, IL-21, STAT’s

## Abstract

Systemic Lupus Erythematous (SLE) is a chronic autoimmune disorder, broadly characterized by systemic inflammation along with heterogeneous clinical manifestations, severe morbidity, moribund organ failure and eventual mortality. In our study, SLE patients displayed higher percentage of activated, inflamed and hyper polarized CD8^+^ T cells, dysregulated CD8^+^ T cell differentiation, significantly elevated serum inflammatory cytokines, higher accumulation of cellular ROS when compared to healthy controls. Importantly, these hyper inflammatory/hyper polarized CD8^+^ T cells responded better to an anti-oxidant than to an oxidant. Terminally differentiated Tc1 cells also showed plasticity upon Oxidant/antioxidant treatment but was in contrast to the SLE CD8^+^ T cell response. Our studies suggest that the differential phenotype and redox response of SLE CD8^+^ T cells and Tc1 cells could be attributed to their cytokine environs during their respective differentiation and eventual activation environs. Polarisation of Tc1 cells with IL-21 drove hyper cytoxicity without hyper polarisation suggesting that SLE inflammatory environment could drive the extreme aberrancy in SLE CD8^+^ T cell.

## 1. Introduction

Systemic Lupus Erythematous (SLE) is a chronic autoimmune disorder, broadly characterized by systemic inflammation along with heterogeneous clinical manifestations. It primarily manifests as a multi organ auto immune disorder, with an inflammatory cytokine *milieu*, exaggerated autoantibody production and dysfunctional immune response eventually leading to life threatening situations [1, 2]. Amongst the multiple immune cells involved in precipitating SLE pathogenesis, T cells play a critical role [3]. Previous reports suggest that the CD4^+^ T cell population in SLE is metabolically compromised and is associated with higher ROS generation, mitochondrial hyperpolarization and reduced glutathione, thus dictating oxidative stress in these patients [4, 5]. However, the role of CD8^+^ T cells in mediating or exacerbating SLE is still unclear and hence our study focusses on defining these cells, attempting if these cells can be plasticized and if these cells can be generated *ex vivo* [6].

The seminal role of CD8^+^ T cells have been well studied and established in intracellular infections and in cancers, where a robust pro-inflammatory response is crucial for clearance of pathogens and cancer. While cytotoxic T cell response with adequate help from helper T cells is exquisitely controlled, uncontrolled pro-inflammatory responses do occur, leading to cytokine storms and cytotoxic protein mediated tissue and organ damage [7]. While the role of CD4^+^ T helper cells is clearer, the role of CD8^+^ T cells in immune pathology and most of the autoimmune diseases is limited, and is primarily focused on Type 1 Diabetes and Multiple Sclerosis [8, 9]. With respect to the status of CD8^+^ T cells in SLE, contradictory reports prevail where specific studies suggest hyper cytotoxicity whereas others indicate hypo cytotoxicity of this population [10–12]. Alongside, recent studies suggest the existence of endophenotypes in SLE, based on the cytotoxic and metabolic profiles of CD8^+^ T cells, while the heterogeneous disease profile is quite clearly also dependent on patient’s intrinsic immune profile, age, disease severity and drug regime [13, 14]. Our studies attempt to profile these inflammatory CD8^+^ T cells in SLE and to examine if cytokines or other factors drive their aberrancy.

Maturation of a naïve CD8^+^ T cell is critically dependent on the cytokine *milieu*, apart from the primary antigen and co stimulus, and is responsible for driving a distinct subtype during their activation. This exclusive and selective cytokine *milieu*, apart from the primary stimulus, co stimulation etc. governs the expression of specific transcription factors, leading to the generation of a specific subtype and inhibition of others. For e.g., the cytokine IL-12 is responsible for inducing the expression of transcription factor, Eomesdermin (Eomes) favoring differentiation of Tc1. Alternatively, cytokine TGF-β and IL-2 is required for differentiation of T suppressor cells i.e., CD8^+^ T cells secreting IL-10 [15,16]. Alongside, its differentiation is also regulated by various signaling molecules such as Reactive Oxygen Species (ROS) and calcium (or Ca^2+^) that are present in the micro environment during CD8^+^ T cell activation. Thus, alteration in the TCR micro environment of these cells can alter phenotypes for e.g., drive an inflammatory towards an anti-inflammatory phenotype or reverse, drive lower than optimal activation, drive immuno suppression, etc,. adversely affecting immune homeostasis and leading to immune pathologies [17, 18].

The present study essentially focuses on understanding the aberrant and inflammatory SLE CD8^+^ T cell phenotype, their oxidant status, role of redox potential in regulating terminally differentiated cytotoxic T cell and potential application in SLE CD8^+^ T cell. Although previous reports discuss the cytotoxic status of SLE CD8^+^ T cells with certain inconsistencies, others suggest an exhausted population. Alongside, there are studies suggesting oxidative stress in SLE CD8^+^ T cells but any direct correlation with either their hyper cytotoxic or exhausted phenotype remains undefined. Consequently, in this study we report predominantly hyper cytotoxic, hyper polarized, higher ROS secreting CD8^+^ T cells, and their oxidant/antioxidant response capabilities. Terminally differentiated human Tc1 cells also exhibited oxidant/antioxidant sensitivity, validating their plastic nature and suggesting that like their CD4^+^ T helper counterparts, cytotoxic CD8^+^ T cells are disease defined and convertible. Alongside, our *ex vivo* studies with respect to human Tc1 cells differentiated in the presence of IL-21 named Tc21, also suggest an increased polarisation towards inflammatory phenotype.

## 2. Materials and Methods

### Clinical characteristics of SLE Patients and demography

For the study, 41 active SLE patients were recruited based on the SLEDAI score. The details of the clinical parameters and medications taken by the patients are given in **Table.1**. In addition, 26 healthy volunteers devoid of any chronic disease, autoimmunity, infection or malignancies, were enrolled for the study. Written Informed consent was taken from all the participants included in the study and Institutional Human Ethics Board, Institute of Life Sciences, approved the study.

### Whole blood and PBMC Phenotyping from SLE patients and healthy controls

100 μl whole blood derived from SLE patients and healthy controls were first incubated with Trustain FcX block (BioLegend) for 30 minutes in the dark, followed by addition of Live/Dead Aqua stain (Invitrogen) for 20 minutes. After the incubation period, monoclonal antibody cocktail of CD3, CD4, CD8, CXCR5 and ICOS was added again followed by incubation for 30 minutes in the dark. Subsequently, RBC lysis was done and the cells were treated with BD Fix and lyse. The cells were then washed at 300g for 8 minutes followed by acquisition in Cytoflex S (Beckman Coulter). To analyze the naïve, effector and memory compartments of CD8^+^ T cells in SLE and HC, CD8^+^ T cells were isolated from PBMC and then stained with CD45RA and CD45RO, followed by acquisition in BD LSR Fortessa (BD Biosciences). The details of reagents and used in the experiments are given in supplementary Table 1. All flow cytometric analyses was done in FlowJo software V10.8 (BD Biosciences) [19].

**Table 1.** Clinical and demographic characteristics of SLE patients and Healthy Controls.

|  | **HC (n=26)** | **SLE (n=41)** |
| --- | --- | --- |
| Age, years, median (IQR) | 28(25-34) | 25(23 – 32) |
| Sex,Female/Male, n(%) | 21(80.76%)/5(19.23%) | 40(97.5%)/1(2.4%) |
| SLEDAI, median (IQR) | N.A | 5(3-6) |
| anti-dsDNA, ELISA, U/mL, median (IQR) | N.D | 35.28 (10.89-74.72) |
| C3, g/L, median (IQR) | N. D | 0.96 (0.64 – 1.25) |
| C4, g/L, median (IQR) | N. D | 0.2((0.19-0.25) |
| CRP, mg/L, median (IQR) | N. D | 7.57(5.78-16.24) |
| History of Nephritis | N. A | 5 |
| **Current drug use** |  |  |
| Prednisolone,n/n, % | N.A | 41/41, 100% |
| Hydroxychloroquine, n/n, % | N.A | 41/41,100% |
| Azathioprine, n/n, % | N.A | 14/41, 34.16% |
| Mycophenolate mofetil,n/n,% | N.A | 24/41, 58.53% |
| Methotrexate,n/n,% | N.A | 12/41, 29.26% |

**SLEDAI- SLE Disease Activity Index, CRP- C-Reactive Protein, C3-Complement Component 3, C4-Complement Component 4, N.D- Not Determined, N.A-Not Applicable, HC- Healthy Control, SLE- Systemic Lupus Erythematosus.**

### Plasma Cytokine Detection Assay

Neat plasma from SLE and healthy controls were run in duplicates to measure 20 T cell cytokines using human Milliplex map cytokine assay kit (Millipore, Billerica, MA). The samples were acquired in a Bio-Plex 200 system (Bio-Rad, Hercules, CA) and cytokine concentrations were calculated using Bio-Plex manager software with a five-parameter (5PL) curve-fitting algorithm applied for standard curve calculation [20].

### Differentiation of CD8^+^ T cells to Tc1 or Tc21 cells from healthy controls

5ml blood was collected from healthy human volunteers and PBMC isolated through Ficoll dependent density gradient centrifugation. CD8^+^ T cells were then isolated from PBMC by negative selection using Dyna beads (Invitrogen, MA, USA) according to the manufacturer’s protocol. Cell purity was checked by flow cytometry and was ascertained to be around 85∼90% consistently. Isolated cells were cultured in RPMI 1640, supplemented with 10% fetal bovine serum, Australian origin (PAN-Biotech), 100 U/ml penicillin, 100 μg/ml streptomycin and 50 mM 2β-mercaptoethanol (Sigma). For Tc1 differentiation, 1 × 10^6^ CD8^+^ T cells were activated by Dynabeads Human T cell activator CD3/CD28 (Gibco, Dublin, Ireland) according to manufacturer’s protocol. Additionally, IL-12 (10 ng/mL), IL-2 (100 IU/ml) and anti-IL-4 (10 mg/mL) was added after 24 hours. For Tc21 differentiation IL-21(25ng/ml) was added along with the above conditions. After 10 days of activation, the cells were then washed with RPMI 1640 media and used for subsequent experiments as indicated [21, 22].

### Cellular and Mitochondrial ROS from Healthy controls and SLE CD8^+^ T cells

Briefly, isolated CD8^+^ T cells were considered for determination of cellular ROS level. A total of 5×10^5^ cells per ml were stained with 500 nM CellROX Deep Red Reagent (Invitrogen) and incubated at 37°C for 60 minutes. 1 mM of 1 μl of Sytox Blue was added during the last 15 minutes of the incubation, to discriminate between live and dead cells. After the incubation, cells were acquired on Cytoflex S (Beckman Coulter). To determine mitochondrial hyperpolarization, a total of 1× 10^6^ cells/ml were stained with 100nM Mitotracker Red CMXRos (Invitrogen) for 15 minutes at 37° C Cells were then washed and acquired on BD LSR Fortessa using a 561nm laser for excitation and 610/20nm emission filter [14, 23].

### Oxidant and antioxidant treatment of CD8^+^ T cells

In brief, 1× 10^6^ cells/ml of CD8^+^ T cells were seeded in flat bottomed 24 well plates and was stimulated with 20ng/ml of PMA (Phorbol 12-myristate 13-acetate, Sigma) and 1μg/ml Ionomycin (Sigma). For ROS induction, 2.5 μM Menadione was added after 30 minutes of PMA/Ionomycin treatment. For quenching of ROS, 5 mM NAC(*N*-Acetyl-L-cysteine) was added to Menadione-treated cells after 15 minutes of Menadione treatment. For assessment of cytokines in the above conditions, 10μg/ml of BFA (Brefeldin A) was added 2.5 hours after PMA/Ionomycin treatment [17, 24].

### Intracellular Staining for Cytokines and Transcription factors

Around 1 × 10^6^ cells/ml were reactivated with 50 ng/ml PMA and 1 μg/ml Ionomycin for 6 h for detecting intracellular cytokines and transcription factor. 10 μg/ml Brefeldin A was added to the culture during the last 3 hours and dead cells were excluded using Zombie Fixable NIR or Aqua Dye Kit (BioLegend, USA). Intracellular cytokine staining was performed using Cytofix/Cytoperm Fixation/Permeabilization Solution Kit (BD Biosciences, San Jose, CA, USA), while transcription factor staining was performed using FOXP3 staining buffer set (eBioscience, San Diego, CA, USA), according to the manufacturer’s instructions [17].

### Phospho-STAT staining for Flow Cytometry

For intracellular staining of human phospho-STAT3/4, unstimulated or stimulated CD8^+^ T cells were fixed with formaldehyde (final concentration of 1.5%) for 30 min at room temperature. They were then permeabilized with ice-cold methanol with vigorous vertexing and incubated at 4° C for 24hr. After washing with staining media (PBS with 1% BSA), cells were stained with respective antibodies for 1 hour at room temperature and samples acquired on BD LSR Fortessa.[17, 25].

### Phospho-mTOR staining for Flow Cytometry

CD8 T cells from SLE patients were treated with menadione or menadione and NAC for 30mins at 37 □followed by activation with PMA/Ion for 10mins.The cells were fixed by adding 1.5% formaldehyde for 30 mins at room temperature. Then they were permeabilised with ice cold methanol with vigorous vortexing and incubated at 4° C for 15mins.After washing with staining media cells were stained with p-mTOR (Ser2448) Antibody for 30 mins at room temperature and then acquired on BD LSR Fortessa[24, 26].

### Statistics

All flow cytometric analysis was done in FlowJo software V10.8 (BD Biosciences). Statistical analysis was performed using the GraphPad Prism software, version 9.0.1. Data is presented as Mean ± Standard Deviation of Mean (SEM). One-way ANOVA with *post hoc* Tukey’s multiple comparison test were used to compare statistics among three groups or more. Mann-Whitney U test was used to compare statistics between HC and SLE patients including cytokine profile from multiplexing. Paired t test was used to compare Tc1 and Tc21 cells from same donors. *P* values less than 0.05 were considered significant (\**P* < 0.05, \*\**P* < 0.01, \*\*\**P* < 0.001, \*\*\*\**P* < 0.0001).

## 2. Results

### SLE CD8^+^ T cells display higher CXCR5 and ICOS and altered compartments

Our primary objective was to examine for altered CD8^+^ T cell populations between healthy volunteers and SLE patients and to report significant differences between them **(Fig 1. A-B**). When we analyzed for the expression of ICOS, a known co-stimulator of CD8^+^ T cells, we found its expression to be significantly higher in SLE patients strongly indicating their higher activation **(Fig 1. C-D)**. Alongside, SLE CD8^+^ T cell expressed higher CXCR5, a chemokine receptor for CXCL13 and known to be functionally responsible for B cell maturation **(Fig 1. E-F)**. The elevated CXCR5 expression in SLE CD8^+^ T cells suggested their resemblance to Tfh cells and is also functionally evinced in the disease where multi organ auto antibodies are encountered apart from inflammation. Altogether, the augmented expression of ICOS and CXCR5 on CD8^+^ T cells of SLE suggested towards a pathogenic CD8^+^ T cell population displaying exaggerated cytotoxicity and indirectly capable of inducing higher antibody response via B cell activation.

**Fig 1.**
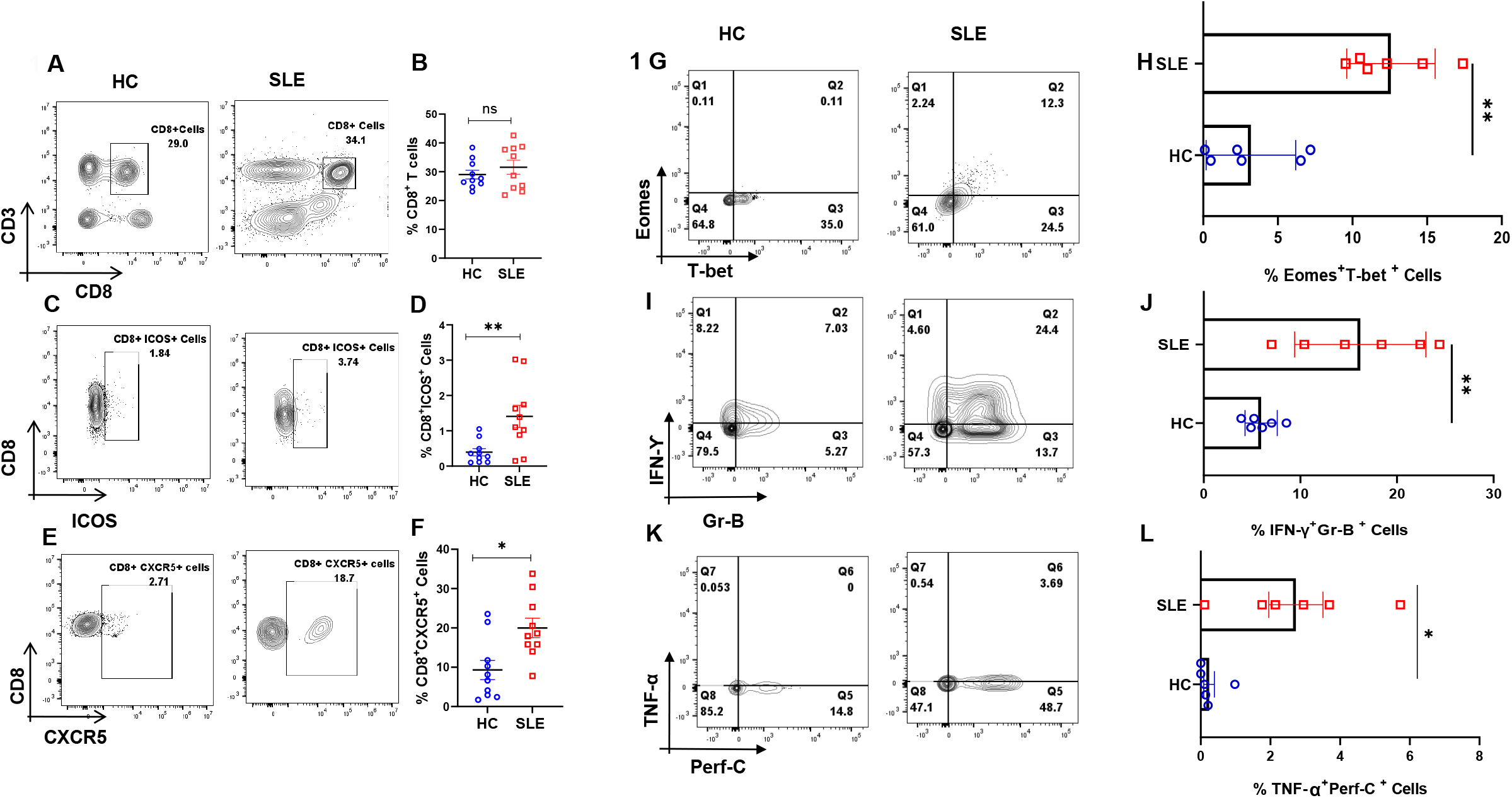
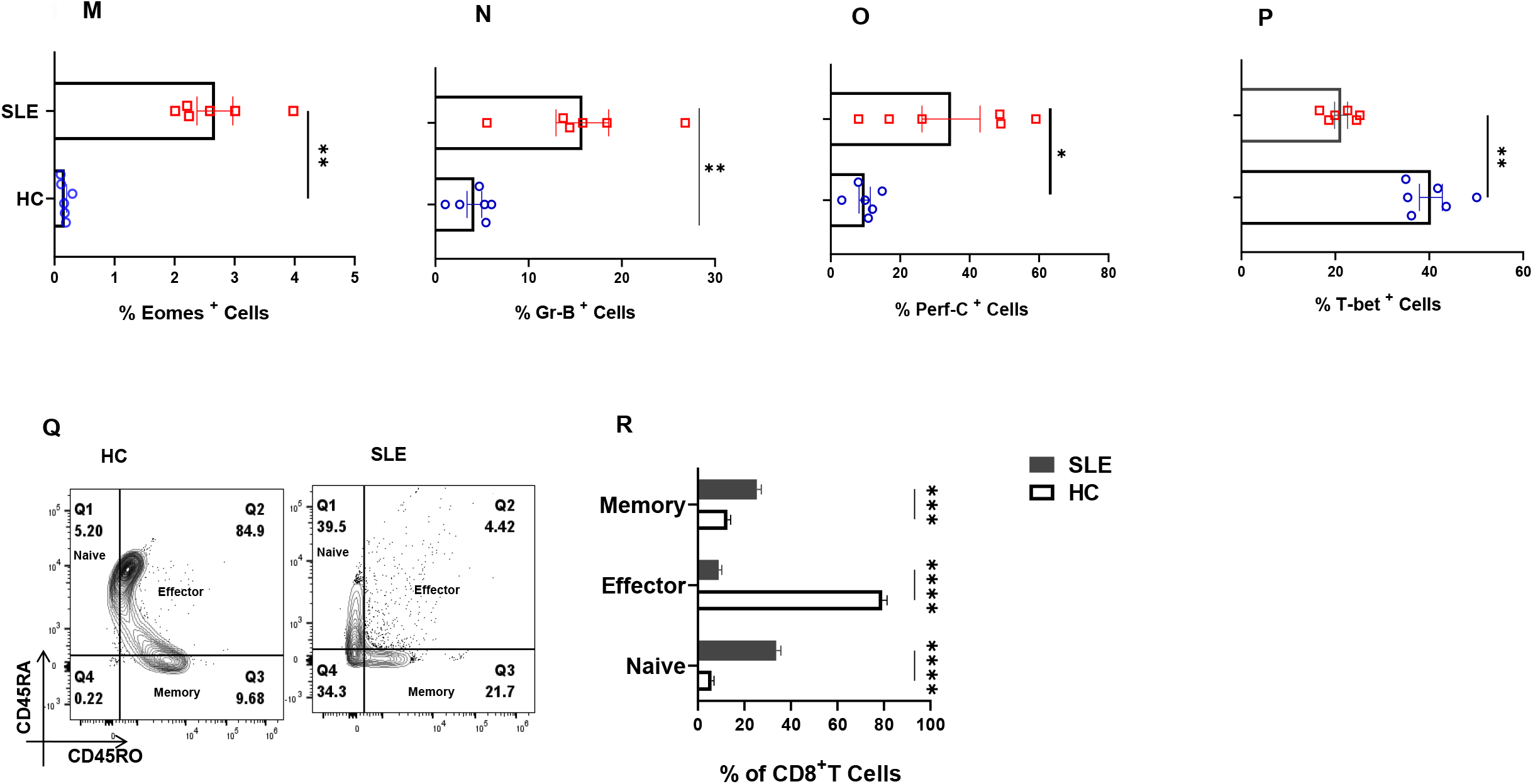
Dysregulated cellular differentiation and expression of activation marker in CD8^+^ T cells of SLE patients. SLE patients display higher activation marker including ICOS and CXCR5 in whole blood. Representative flow cytometry plots show % CD8^+^ T cells in whole blood of healthy controls (n=10) and SLE patients (n=10) **(A)** and cumulative graphical representation **(B)**. Representative plots showing % ICOS expression on CD8^+^ T cells from whole blood of HC (n=10) and SLE patients (n=10) (**C)** and cumulative graphical representation **(D)**. Representative plots showing expression of % CXCR5 expression on CD8^+^ T cells from whole blood of HC (n=10) and SLE patients (n=10) (**E)** and cumulative graphical representation **(F)**. SLE patients (n=6) display higher expression of Eomes and T-bet dual positive CD8^+^ T cells than healthy controls (n=6) while a reversed trend is observed for T-bet, representative flow cytometry plots **(G)** and cumulative graphical representation **(H**). Representative flow cytometry plots showing increase in inflammatory cytokines IFN-γ, TNF-α and cytotoxic molecules Granzyme-B and Perforin-C in SLE patients (n=6) as compared to healthy control (n=6) **(I, K)** and cumulative graphical representation **(J, L)**. Single positive Eomes **(M)**, Gr B **(N)**, Perf C **(O)** and T-bet **(P)** are also shown from SLE CD8^+^ T cell. Representative plot showing naive, effector, memory compartments between healthy controls (n=10) and SLE patients (n=10) (**Q)** and cumulative graphical representation **(R)**. Error bar indicates SEM. Mann-Whitney U Test has been used to compare between the groups. 2-way ANOVA was used to compare two variables among more than two groups. p<0.05 was considered statistically significant (*), p<0.01 was considered to be very significant (**), *P* < 0.001 was considered to highly significant (***), *P* < 0.0001 was considered extremely significant (****) ns, not significant.

Since SLE is characterized by very high systemic inflammation and our initial characterisation of the SLE CD8^+^ T cell population showed aberrancy, we then examined for known inflammatory cytokines such as IFN-γ and TNF-α, cytotoxic molecules Granzyme B and Perforin C, and transcription factors T-bet and Eomes. Here, we observed that co expression of IFN-γ-Granzyme-B and TNF-α-Perforin C were higher in SLE CD8^+^ T cells **(Fig 1. I-L)** as was the dual expression of transcription factors T-bet and Eomes **(Fig 1. G-H)**. Interestingly, the expression of Eomes, Gr B and Perf C **(Fig 1. M-O)** was significantly higher in SLE CD8^+^ T cells while single positive T-bet **(Fig 1.P)** was lower in SLE and IL-10 did not show any significant change (Data not shown). The above results indicate that SLE CD8^+^ T cells displayed significantly higher cytotoxic and inflammatory phenotype as compared to controls. Interestingly, when we compared between compartments, SLE CD8^+^ T cells had higher percentages of naïve and memory cells **(Fig 1. Q, R)** suggesting a strong recall response. In addition, we found a higher percentage of memory and naïve CD8^+^ population in SLE probably as a consequence of higher levels of one of its driving cytokines, IL-15, suggesting a possible role of IL-15 during systemic inflammation.

### SLE patients display higher pro-inflammatory cytokine profile

To understand systemic inflammation in SLE patients, we assessed for 20 T cell cytokines and found that most of the pro-inflammatory ones were elevated in the plasma of SLE patients **(Fig 2. A)** and in direct contrast, only IL-13 amongst the anti-inflammatory was elevated **(Fig 2. B)**. Amongst the pro-inflammatory cytokines that promote Type I response, IFN-γ, IL-12p40, TNF-α were significantly elevated in SLE and additionally IL-6, IL-1β, IL-17A, IL-17E and IL-15 were also elevated **(Fig 2. A)**. Most of the anti-inflammatory cytokines such as IL-4, IL-10, IL-5, IL-9 did not show any significant difference in SLE patients as compared to healthy controls **(Fig 2. B)**. Altogether, this suggested that despite a strong inflammatory reaction in these patients, the anti-inflammatory response was markedly inadequate or even non-functional. We infer that the significantly higher levels of inflammatory cytokines can profoundly activate cells of both innate and adaptive response creating a positive feedback loop, in turn exacerbating the pathology of SLE. Most importantly our results envisage an highly unpredictable and dysregulated immune response due to the diversity and flexibility of the cytokine mix.

**Fig 2.**
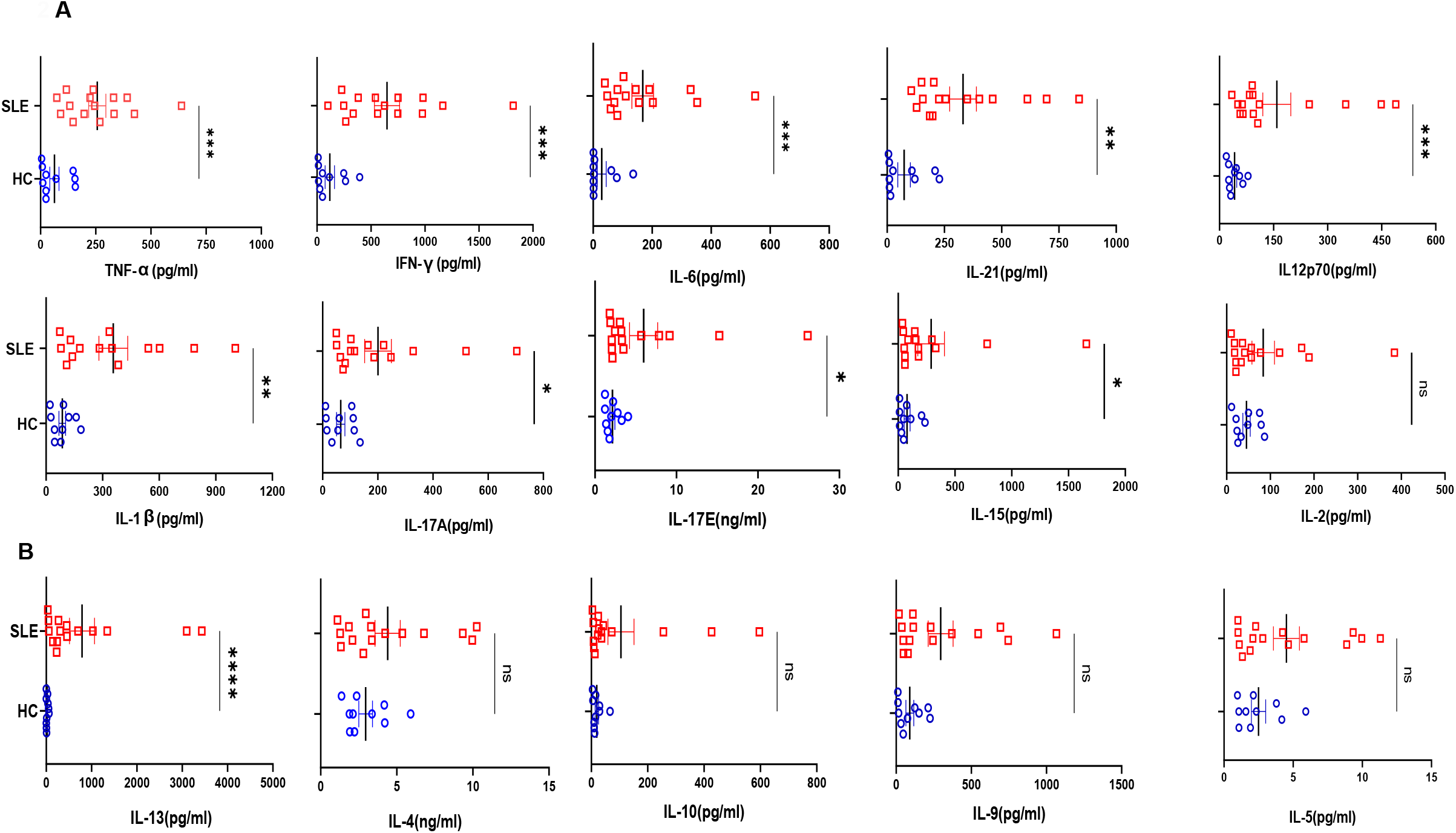
Cytokines analyses in SLE patients and healthy controls. Representative figures from 20 T cell cytokines analyzed in SLE patients’ plasma (n=15) and healthy controls (n=10) are represented as plunge plots. Inflammatory cytokines including IFN-γ, IL-12p40, TNF-α, IL-6, IL-1β, IL-17A, IL-17E, IL-21, IL-15 and IL-21 were significantly elevated in SLE patients as compared to HC **(A)**. Amongst anti-inflammatory cytokines only IL-13 was elevated in SLE patients while all others including IL-10, IL-4, IL-5, IL-9 did not show any difference between the two groups **(B)**. Error bar indicates SEM. Mann-Whitney U Test was performed to compare between the two groups, p<0.05 was considered statistically significant (*), p<0.01 was considered to be very significant (**), *P* < 0.001 was considered to highly significant (***), *P* < 0.0001 was considered extremely significant (****) ns, not significant.

### SLE CD8^+^ T cells display higher ROS and mitochondrial hyperpolarization

Next, we examined the levels of cellular Reactive Oxygen Species (cROS) in SLE CD8^+^ T cells as ROS is known to be generated during TCR activation and signaling and has been shown to fine tune the balance between pro and anti-inflammatory response. ROS maintains metabolic fitness of T cells and excessive accumulation of ROS is known to cause to adversely impact immune responses. In this study, we found increased ROS generation in SLE CD8^+^ T cells **(Fig 3. A-C)** that is in line with previous reports suggesting that ROS was higher in SLE lymphocytes and that the antioxidant, glutathione was reduced in these cells. In addition, earlier studies have shown that CD4^+^ T cells in SLE have mitochondrial abnormalities including increased mitochondrial size and membrane disruption and led us to examine for altered mitochondrial status and ROS levels of SLE CD8^+^ T cells. Here, we observed that the mitochondria was also hyperpolarised that could have resulted in the increased ROS production or *vice versa*, and consequently or subsequently could have led to altered redox status in these cells **(Fig 3. D-F)**. Altogether, mitochondrial hyperpolarization and consequent augmented ROS generation and in SLE CD8^+^ T cells suggested a significantly dysregulated redox gradient in them.

**Fig 3.**
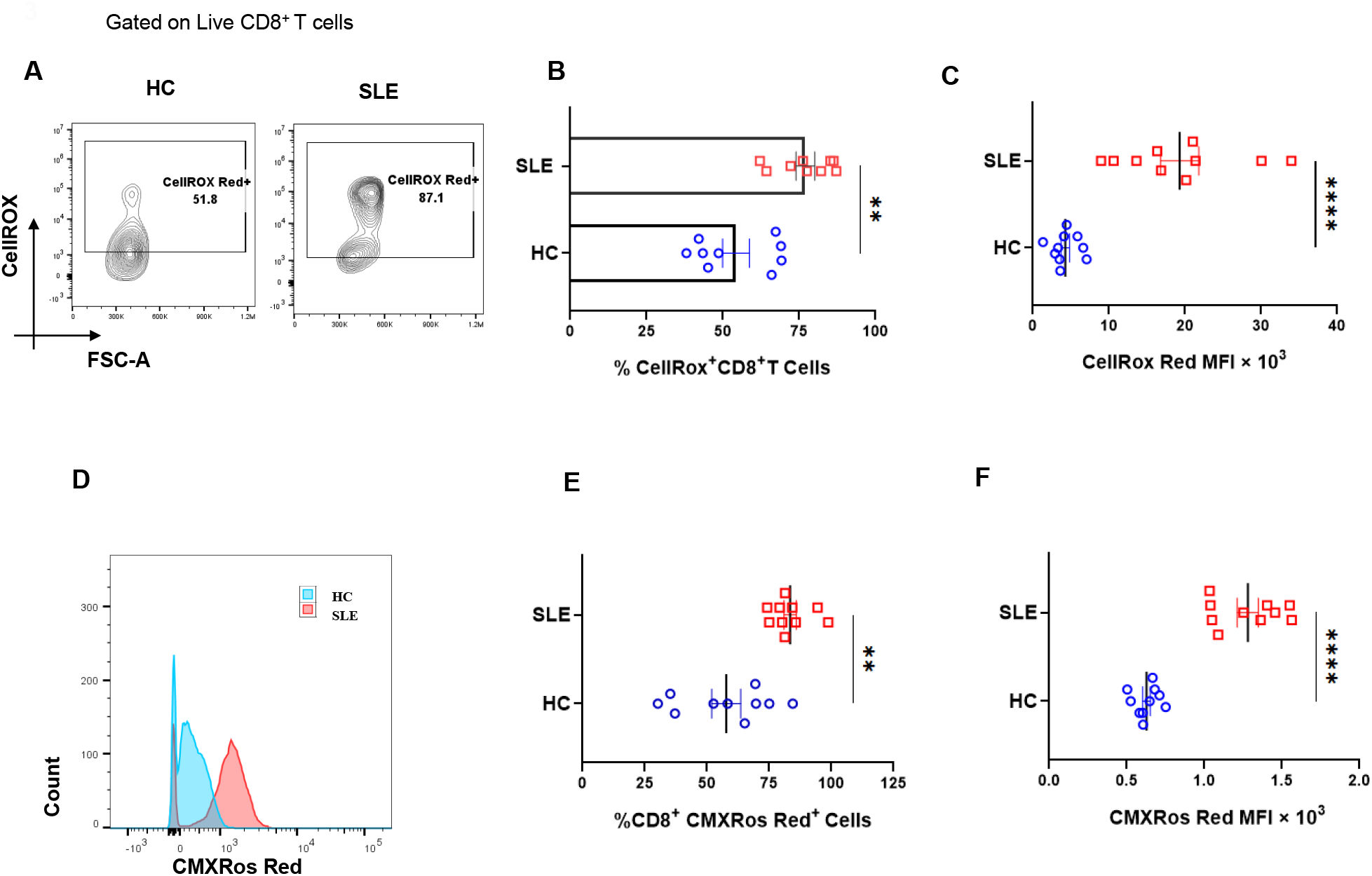
Cellular ROS (cROS) and mitochondrial hyperpolarization in SLE CD8^+^ T cells. CD8^+^ T cells from SLE displayed higher cellular ROS than Healthy Controls. Representative flow cytometry plots showing CD8^+^ T cells from HC (n=10) and SLE (n=10) **(A)** stained with CellROX Red to determine cellular ROS in CD8^+^ T cells and cumulative graphical representation of the % cells expressing CellROX **(B)**. Graphical representation showing difference in Median fluorescence Intensity (MFI) of CellROX between HC and SLE **(C)**. Representative flow cytometry plots showing CD8^+^ T cells from HC (n=10) and SLE (n=10) stained with CMXROS Red to determine mitochondrial hyperpolarisation **(D)**. Cumulative Graphical representation of the % cells expressing CMXROS Red **(E)** and Graphical representation showing difference in Median fluorescence Intensity (MFI) of CMXROSRed between HC and SLE **(F)**. Error bar indicates SEM. Mann-Whitney U Test has been used to compare between the groups. p<0.05 was considered statistically significant (*), p<0.01 was considered to be very significant (**), *P* < 0.001 was considered to highly significant (***), *P* < 0.0001 was considered extremely significant (****) ns, not significant.

### Redox controls pro- and anti-inflammatory markers in SLE CD8^+^ T cells

Our mitochondrial and ROS status studies in SLE showed significant aberrancy and strongly suggested to examine their pro and anti-inflammatory response characteristics. When SLE CD8^+^ T cells were treated with a known Oxidant (Menadione) or anti-oxidant (NAC), we found that SLE CD8^+^ T cells responded better to NAC with significant reduction in IFN-γ production **(Fig 4. A, G)**. On the contrary, we did not find any difference in the expression of IL-10 **(Fig 4. A, F)** among untreated cells, cells treated with menadione and or cells treated with both menadione and NAC. We also report a significant decrease in the expression of Eomes and TNF-α in NAC treated SLE CD8^+^ T cells when compared to Menadione treated cells **(Fig 4. D, E)** and with the PMA/Ion treated population. This is in corroboration with previous studies, where it has been shown that NAC inhibits TNF-α in cells exhibiting higher oxidative stress **(Fig 4. E, I)** and suggested that the restoration of anti-oxidant potential in the CD8^+^ T cells in SLE reverses the inflammatory phenotype of the CD8^+^ T cells in SLE.

**Fig 4.**
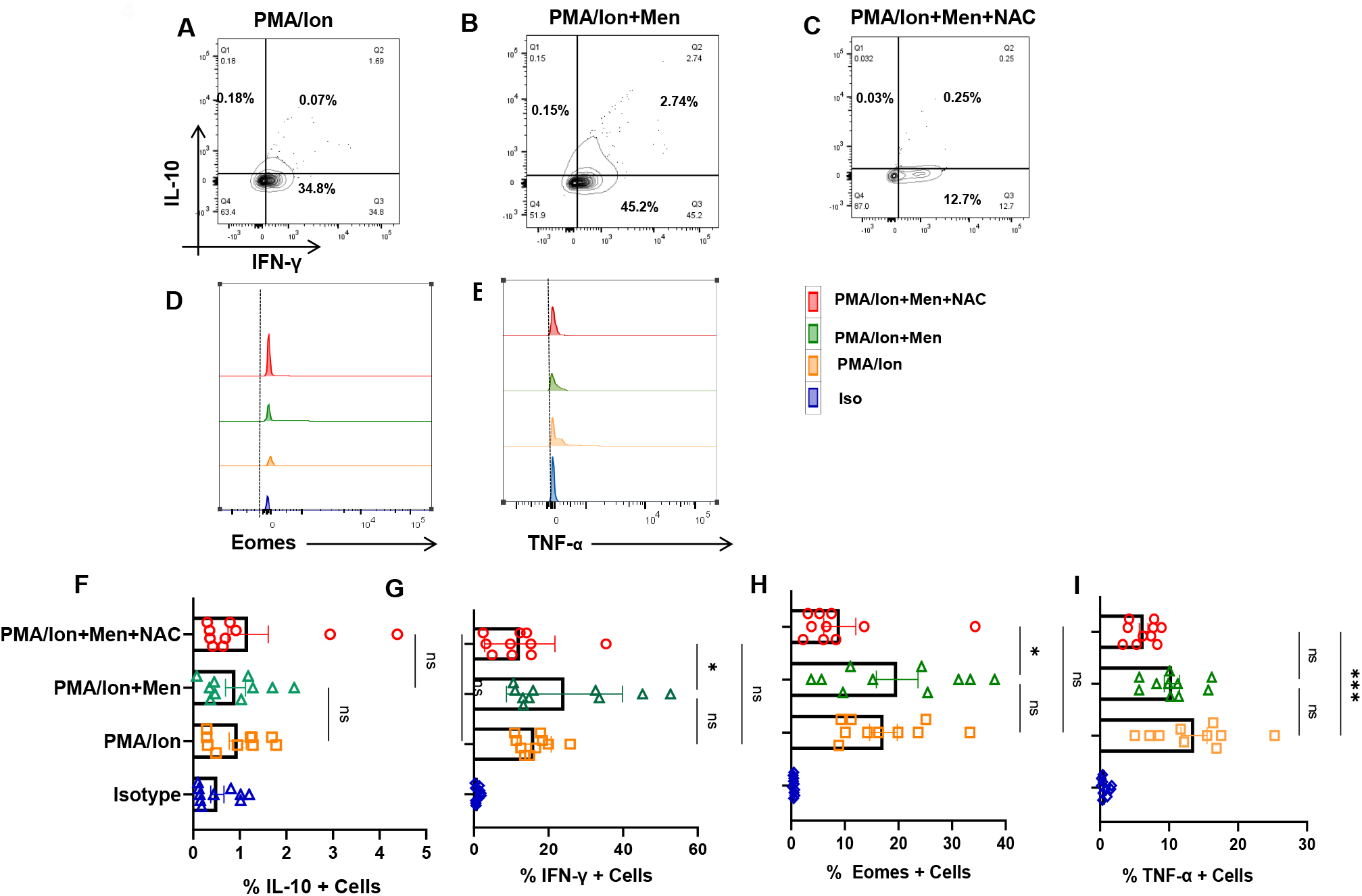
Redox modulation of inflammatory cytokines and transcription factor from SLE CD8^+^ T cells. CD8^+^ T cells were isolated from PBMC of SLE patients and treated with 2.5μM Menadione with or without NAC for 4 hrs. and processed according to the protocol mentioned in the method section. Representative figures showing reduction in IFN-γ, TNF-α and Eomes following NAC (5mM) treatment as compared to their activation by PMA/Ion or treatment with menadione (2.5μM) **(A-E)**. Cumulative graphical representation showing changes in IL-10 and IFN-γ after Menadione and NAC treatment **(F, G)**. Cumulative graphical representation showing changes in Eomes and TNF-α after Menadione and NAC treatment **(H, I)**. Error bar indicates SEM. The experiments were done from 20 SLE patients and all samples were tested for significance by using one-way ANOVA followed by Tukey’s multiple comparison. p<0.05 was considered statistically significant (*), p<0.01 was considered to be very significant (**), *P* < 0.001 was considered to highly significant (***), *P* < 0.0001 was considered extremely significant (****) ns, not significant.

### Modulating ROS reduces p-STAT4 and p-mTOR from SLE CD8^+^ T cells

ROS is known to upregulate p-STAT3 and consequently led us to examine for the levels of p-STAT’s including p-STAT4 in SLE CD8^+^ T cells. We report no significant change in p-STAT3 levels from SLE CD8^+^ T cells **(Fig 5. A, D)** but evinced that NAC was able to downregulate activated p-STAT4 levels in SLE CD8^+^ T cells **(Fig 5. B, E)**. Our studies indicated that hyper cytotoxic/polarized SLE CD8^+^ T cells display p-STAT3 that was insensitive to any Oxidant/Antioxidant treatment while p-STAT4 was anti-oxidant sensitive. Further, this selective sensitivity could be as a result of the inflammatory cytokine environs that SLE CD8^+^ T cells exist or encounter during activation. Crucially, p-STAT3’s insensitivity could be due to the complete uncoupling or desensitization of the IL-10 pathway, and the subsequent inability to modulate IL-10 secretion could be due to the persistent inflammatory environment **(Fig 4. A, F)**. On the other hand, modulation of p-STAT4’s activity with NAC again points to an inflammatory cytokine signaling pathway still responsive to anti-oxidants. Since mitochondrial hyperpolarization activates mammalian target of Rapamycin (mTOR) which can modulate T cell differentiation and cell death pathways, we analyzed for their oxidant/anti-oxidant sensitivity in SLE CD8^+^ T cells. p-mTOR from SLE CD8^+^ T cells again was significantly NAC sensitive **(Fig 5. C, F)** consolidating our earlier conclusion and maybe suggesting upstream presence to p-STAT3 and 4. These results also suggested that p-mTOR could be modulated in SLE CD8^+^ T cells to counter their inflammatory potential. However, p-mTOR’s response with Menadione or NAC suggests no correlation with IL-10, while IFN-γ and Eomes had direct co relate with NAC.

**Fig 5.**
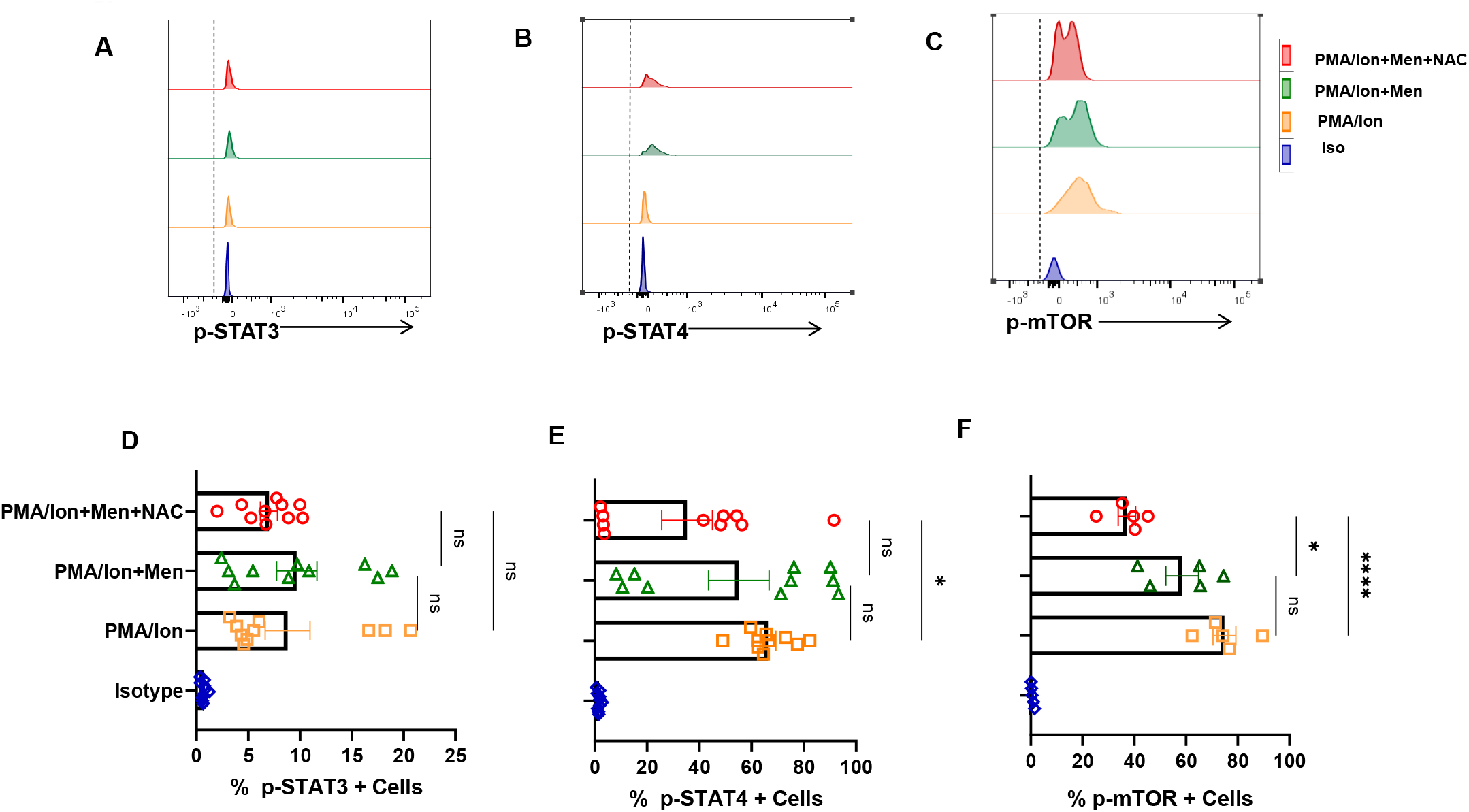
Antioxidant alters expression of p-STAT4 and p-mTOR from SLE CD8^+^ T cells. CD8^+^ T cells were isolated from PBMC of SLE patients and treated with 2.5μM Menadione with or without NAC for 4 hrs. and processed according to the protocol mentioned in the method section. Representative figures showing reduction in p-STAT4 and p-mTOR following NAC (5mM) treatment as compared to their activation by PMA/Ion or treatment with Menadione (2.5μM**) (B-C)**. Cumulative graphical representation showing changes in p-STAT3, p-STAT4 and p-mTOR after menadione and NAC treatment **(D-F)**. Error bar indicates SEM. The experiments were done from 15 SLE patients (n=10) for p-STAT assay and (n=5) for p-mTOR assay and all samples were tested for significance by using one-way ANOVA followed by Tukey’s multiple comparison. p<0.05 was considered statistically significant (*), p<0.01 was considered to be very significant (**), *P* < 0.001 was considered to highly significant (***), *P* < 0.0001 was considered extremely significant (****) ns, not significant.

### Effector Tc1 cells exhibit ROS sensitivity

In order to confirm that SLE CD8 T cell response was indeed characteristic of an inflammatory cytotoxic T cell, that modulating ROS was indeed capable of regulating the inflammatory/anti-inflammatory response, and that the above results were not artifacts, we treated *ex vivo* differentiated human Tc1 cells to oxidant/antioxidant reagents. Differentiated Tc1 cells displayed significant IFN-γ production and extremely low IL-10 expression with PMA/Ion activation and reversed the trend with Menadione treatment **(Fig 6. A, B)**. Not surprisingly, the expression of both IFN-γ and IL-10 was restored upon treatment with anti-oxidant NAC **(Fig 6. C)**. Most importantly, our results were similar amongst the tested batches of terminally differentiated Tc1 cells **(Fig 6. I-J)** where addition of exogenous ROS to Tc1 cells drives an anti-inflammatory response, and reversed with an anti-oxidant. Additionally, Eomesdermin (Eomes) as one of the crucial transcription factors, and TNF-α as one of the crucial pleiotropic cytokines that are required to sustain the cytotoxic and inflammatory response of CD8^+^ T cells were also examined. When we tested for these proteins in terminally differentiated Tc1 cells with PMA/Ion, we observed significant expression of Eomes^+^, TNF-α^+^ and double positive populations **(Fig 6. E)**. Upon Menadione treatment, percentages of both Eomes positive and dual positive Eomes and TNF-α population decreased **(Fig 6. F)** suggesting their oxidant sensitivity **(Fig 6. K-L)**. The above results confirmed that both terminally differentiated Tc1 cells as well as SLE CD8^+^ T cells were Redox sensitive, but were contra indicated in the latter due to the highly inflammatory disease conditions.

**Fig 6.**
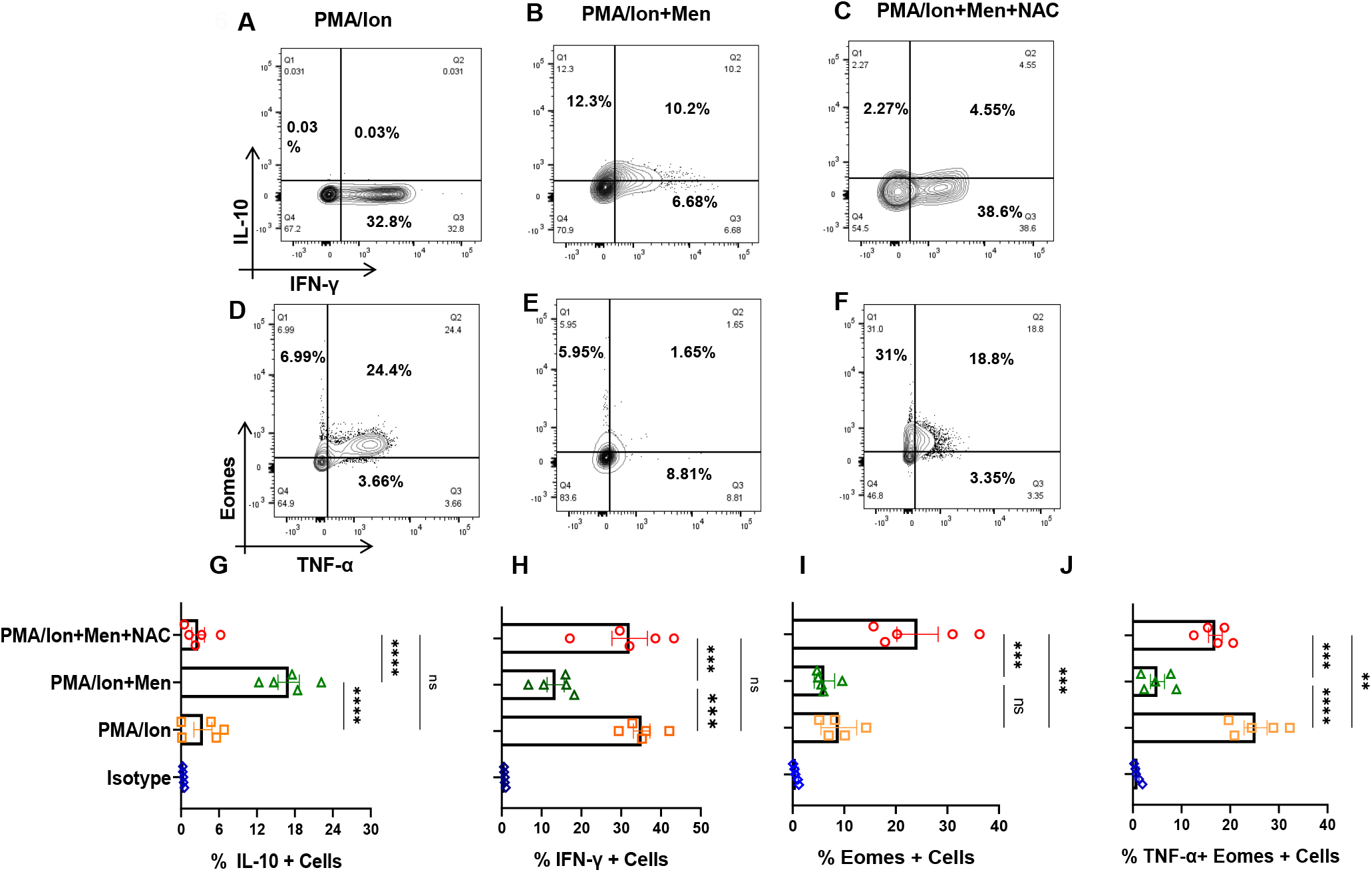
Tc1 cell cytokines are Redox sensitive. CD8^+^ T cells were isolated from PBMC and differentiated for 10 days following which they are reactivated with PMA/Ion for 4 hrs. And stained with desired antibodies as described in materials and method section Representative figures showing reduction in IFN-γ and increase in IL-10 production following Menadione (2.5μM) treatment as compared to their activation by PMA/Ion which is again getting restored following treatment with 5Mm NAC **(A-C)**. Representative figures showing reduction in TNF-α and Eomes expression following Menadione (2.5μM) treatment as compared to their activation by PMA/Ion which is again getting restored following treatment with 5Mm NAC **(D-F)**. Cumulative graphical representation showing significant increase in IL-10 following ROS induction **(H)**. Cumulative graphical representation showing significant decrease in IFN-γ following ROS induction **(I)**. Cumulative graphical representation showing decrease in the expression of Eomes with Menadione treatment which was restored by subsequent NAC treatment **(J)**. Cumulative graphical representation showing reduction in Eomes, TNF-α double positive Tc1 cells with Menadione treatment which was restored by subsequent NAC treatment **(K)**. Error bar indicates SEM. All samples were tested for significance by using one-way ANOVA followed by Tukey’s multiple comparison. p<0.05 was considered statistically significant (*), p<0.01 was considered to be very significant (**), *P* < 0.001 was considered to highly significant (***), *P* < 0.0001 was considered extremely significant (****) ns, not significant.

### ROS induces expression of p-STAT3 from Tc1 cells

We furthered our studies with Tc1 cells to examine for their JAK-STAT pathway perturbations under oxidative stress condition that is known to lead to disruption of cellular homeostasis. Therefore, we analyzed for the phosphorylation status of STAT molecules, when cytokine production was altered through ROS induction. Here we found that, following ROS induction through menadione, the switch towards anti-inflammatory cytokine production is accompanied by increase in p-STAT3 and reduction in p-STAT4 **(Fig 7. B, E**) where it is known that p-STAT3 is known to be required for IL-10 whereas p-STAT4 is required for IFN-γ production. The trend was reversed with antioxidant NAC, causing an increase of p-STAT4 and decrease in p-STAT3 **(Fig 7. C, F)** and was statistically similar in all the batches tested **(Fig 7. G, H)**. Therefore, our data suggested that ROS regulated the pro and anti-inflammatory cytokine production by affecting the phosphorylation status of respective STAT molecules in *ex vivo* differentiated human Tc1 cells. p-STAT3’s modulation in terminally differentiated Tc1 cells gave credence to our hypothesis that hyper inflammatory cytokine status could desensitize p-STAT3 in SLE CD8^+^ T cells.

**Fig 7.**
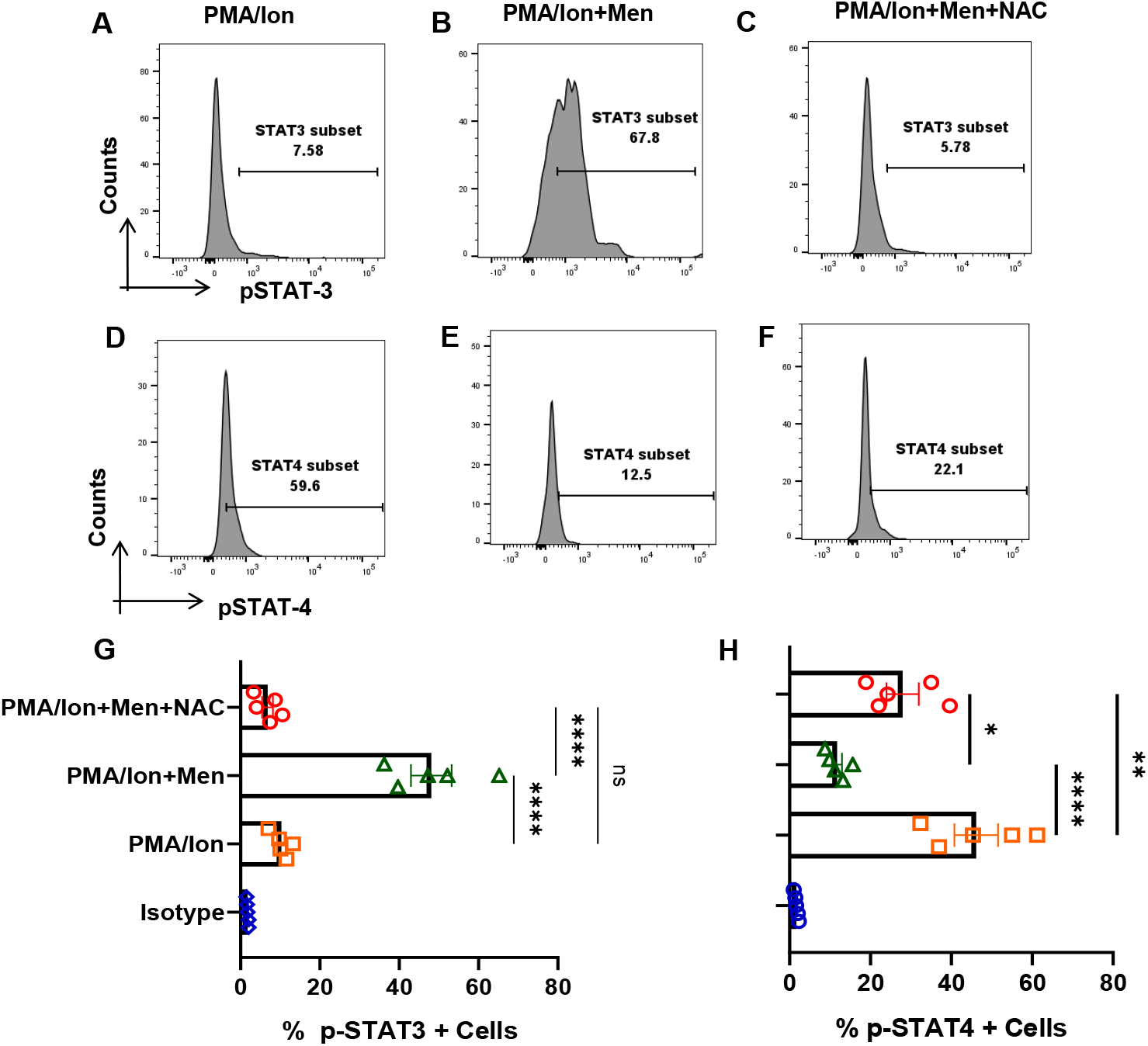
ROS modulates p-STAT’s from terminally differentiated Tc1 cells. Tc1 cells were stimulated with PMA/Ion and treated with 2.5μM Menadione with or without NAC for 3 hrs. and processed according to the protocol mentioned in the method section. Representative figures show increase in p-STAT3 and decrease in p-STAT4 following induction of ROS through Menadione as compared to their activation by PMA/Ion. Also following treatment with ROS scavenger NAC, the result is reversed **(A-F)**. Cumulative graphical data showing significant increase in p-STAT3 expression following ROS induction and their restoration through NAC **(G)**. Cumulative graphical data showing significant increase in p-STAT4 expression following ROS induction and their restoration through NAC **(H)**. Error bar indicates SEM. The experiments were done from 5 healthy donors and all samples were tested for significance by using one-way ANOVA followed by Tukey’s multiple comparison. p<0.05 was considered statistically significant (*), p<0.01 was considered to be significant, p<0.01 was considered to be very significant (**), *P* < 0.001 was considered to highly significant (***), *P* < 0.0001 was considered extremely significant (****) ns, not significant.

### IL-21 polarizes Tc1 cell towards more inflammatory phenotype

We furthered our investigations into understanding how naïve CD8^+^ T cell differentiated into the hyper cytotoxic and inflammatory SLE CD8^+^ T cell. As we evinced significantly elevated levels of IL-21 from SLE patients’ we hypothesized that IL-21 could play a significant role in driving hyper cytotoxicity as IL-21 is a pleotropic cytokine known to be pathogenic in SLE by inducing Tfh cells and autoantibody production. Therefore, to examine its effect on CD8^+^ T cells, we differentiated human CD8^+^ T cells into conventional Tc1 and Tc1 in the added presence of IL-21 (Tc21) and not surprisingly found the latter, a highly inflammatory phenotype. Significantly higher percentage of Tc21 cells were dual positive for IFN-γ-Gr B and TNF-α-Perf C when compared to terminally differentiated conventional Tc1 cell **(Fig 8. A-D)**. This conclusively proved that IL-21 in SLE could be the major cytokine that drove the hyper cytotoxic and inflammatory SLE CD8^+^ T cell that we encountered with possibly Tfh cells as IL-21’s cellular source. To further substantiate the role of ROS in Tc21 cells we then examined for cellular ROS in *ex vivo* differentiated Tc1 and Tc21 cells. Interestingly we did not find any difference between these two subtypes indicating that these cells were not metabolically compromised **(Fig 8. E-F)**. Most importantly this suggested that IL-21 supplementation alone as in our experimental condition was not sufficient to drive hyper polarization. Whether extended IL-21 exposure alone could alter CD8^+^ T cell mitochondrial properties or the SLE inflammatory cytokine *milieu* and disease duration is mandatory for the transformation will remain a clinical and immunological challenge to determine.

**Fig 8.**
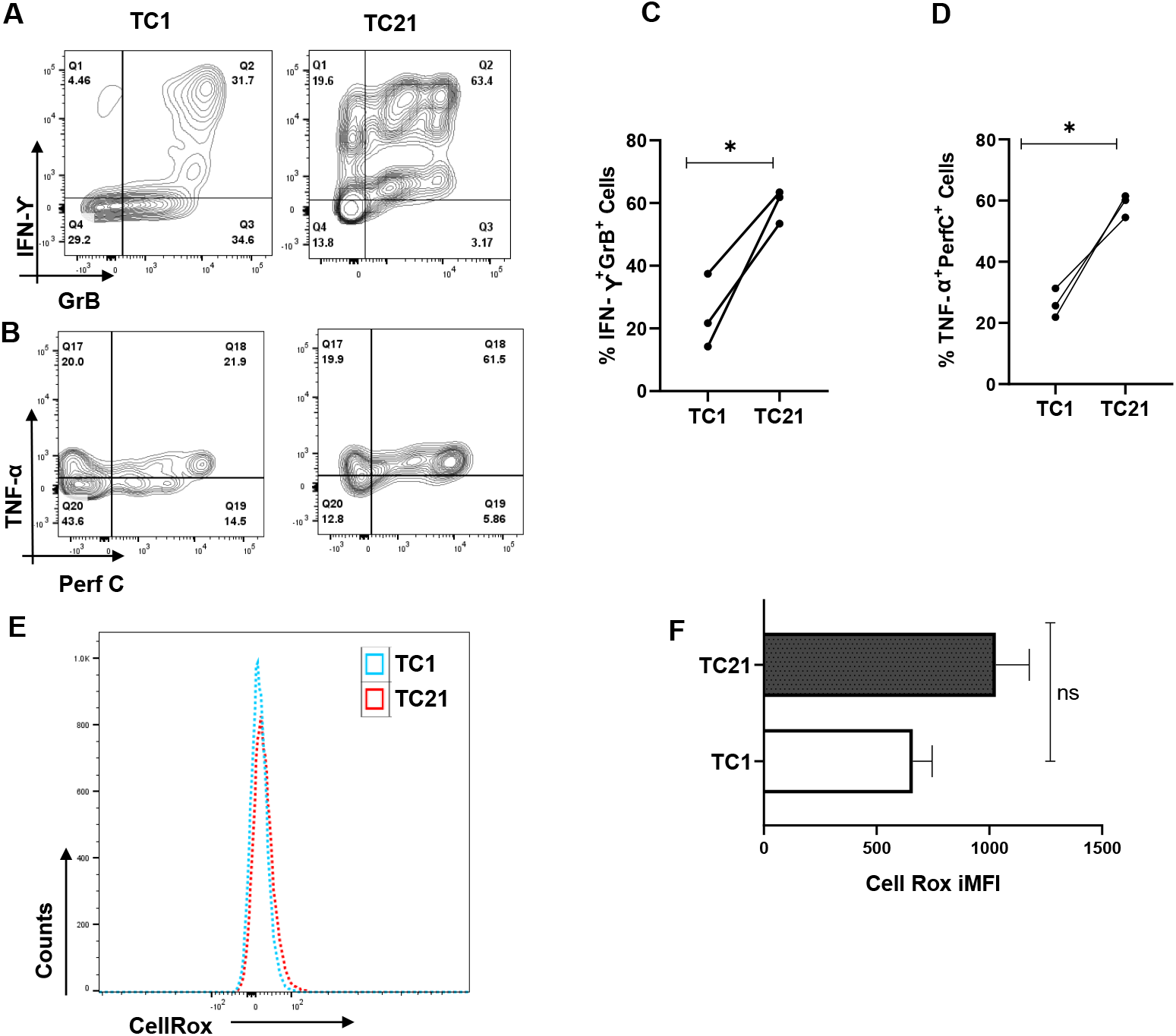
IL-21 drives hyper cytotoxicity in Tc1 cells. CD8^+^ T cells were isolated from healthy controls (n=3) and cultured for 10 days with and without IL-21 and assessed for different cytokines. Representative flow cytometry figures showing increase in dual positive IFN-γ and Granzyme B and dual positive TNF-α and Perf-C when IL-21 is added to the Tc1 culture **(A, B)** and cumulative graphical data showing the same **(C, D)**. Representative figure showing cellular ROS in Tc1 and Tc21 **(E)** and cumulative graphical data showing the same **(F)**. Error bar indicates SEM. All samples were tested for significance by paired t test (two-tailed) for Tc1 and Tc21cells. p<0.05 was considered statistically significant (*), p<0.01 was considered to be significant, p<0.01 was considered to be very significant (**), *P* < 0.001 was considered to highly significant (***), *P* < 0.0001 was considered extremely significant (****) ns, not significant. Mann-Whitney U Test has been used to compare between the groups. p<0.05 was considered statistically significant (*), p<0.01 was considered to be significant, p<0.01 was considered to be very significant (**), *P* < 0.001 was considered to highly significant (***), *P* < 0.0001 was considered extremely significant (****) ns, not significant.

## Discussion

We examined for the role of CD8^+^ T cells in SLE, a known chronic autoimmune disorder that manifests with dysregulated immune response and associated with severe morbidity and mortality without aggressive clinical intervention. The role of aberrant CD4^+^ T cell in SLE has been examined and shown with greater clarity than CD8^+^ T cells and this study clearly establishes the latter’s role in mediating and sustaining inflammation and cytotoxicity, and strongly suggests that they play a seminal role if not being primary in tissue destruction [27–29].

We first profiled for SLE CD8^+^ T cells and report higher expression of activation markers such as ICOS, CXCR5, Eomes and T-bet, inflammatory and cytotoxic molecules such as IFN-γ, TNF-α, Gr-B and Perforin-C [30, 31]. We evinced significantly higher Eomes^+^T-bet^+^, IFN-γ^+^Gr-B^+^ and TNF-α^+^Perf-C^+^ cells in SLE confirming our hypothesis for the role of a “Hyper cytotoxic” CD8^+^T cell phenotype [16] in mediating inflammation. Interestingly we also found that Naïve and Memory compartments were higher in SLE CD8^+^ T cells. While the inflammatory markers lent easy understanding and credence to the disease status, we found the increase in the naïve and memory CD8^+^ T cell compartments to be a bit intriguing and examined the causal cytokines for their elevated levels.

To elaborate on these findings, we did a multiplex analysis of the systemic cytokines present in both healthy volunteers and SLE patients and found that all the inflammatory cytokines of the panel such as IFN-γ, IL-12p40, TNF-α, IL-6, IL-1β, IL-17A, IL-17E and IL-15 were elevated. Apart from that, certain cytokines including IL-12p40, IL-15 and IL-21 that promote memory T cell formation were elevated significantly, strongly suggesting their possible contribute to the higher Memory compartment profile seen in SLE patients and may also be responsible for exacerbating the disease pathology through a recall response [32–34]. Elevated IL-2, IL-12 and IL-15 also suggest that these cytokines could boost naïve T cell genesis and subsequent activation [35]. Importantly, the above cytokines specifically IL-15 signalling is essential for normal immune system functions, to stimulate T cell proliferation and inhibit IL-2-mediated activation-induced cell death. In addition, IL-15 is required for the development, survival, and activation of natural killer (NK) cells, homeostasis of natural killer T (NKT) cells and intraepithelial lymphocytes, and maintenance of naïve and memory CD8^+^ T cells. While the naïve and memory compartment rise in SLE is explained we do recognize that the elevated naïve cells could also be T_scm_ stem cell like memory cells as it is well known that these cells share some overlapping features with the naïve cell population and are reported to be increased in SLE [36]. As we did not analyze for specific markers for stem cell like memory cells, we could not delineate the correlation between naïve cells and stem cell like memory cells further. However, anti-inflammatory cytokines such as IL-4, IL-10, IL-5, IL-9 were higher but did not show any significant difference suggesting imbalance and a dysregulated anti-inflammatory response. But higher than normal levels strongly suggest that these cytokines could be implicated to enhance B cell proliferation, differentiation, and MHC class II expression, regulate B cell immunoglobulin production, enhanced mast cell protease expression, and goblet cell hyperplasia and mucus production [37]. Taken together, these findings suggested that SLE is associated with inflammatory cytotoxic T cells, elevated inflammatory cytokines, cytotoxic proteins and associated cytokines that could drive unwarranted antibody production. All these aberrations of the immune system profoundly affect the progression of the disease-causing morbidity and eventual mortality if not aggressively treated.

Others and we have previously reported the seminal role of ROS in mediating and modulating TCR activation, MAPK activity, cytokine response and aberrant T cell response in immune pathology[17,24]. With respect to the autoimmune disorder SLE, apart from the inflammatory cytokine profile, these patients have oxidative stress in the peripheral blood lymphocytes and have reduced levels of thiols and glutathione reductase that serve as physiological antioxidants[38]. Therefore, to understand oxidative stress occurring in the CD8^+^ T cells of SLE, we examined for cellular ROS and mitochondrial hyperpolarization and report that both were higher. Although our study essentially confirmed accumulated cellular ROS and mitochondrial hyper polarization as statistically significant, we were unable to determine the sequential order between the two. Importantly, our data clearly proved that oxidative stress conditions in SLE CD8^+^ T cells, specifically from the mitochondria, had serious implications with respect to immune homeostasis [37, 38]. Given that SLE had significant inflammatory cytokines elevated and that ROS is implicated in inflammation we directed our studies to try and understanding the above factors and their association [14, 39– 41].

With respect to SLE patients, we observed that CD8^+^ T cells responded better with an antioxidant with significant decrease in IFN-γ, Eomes and TNF-α expression. The above results indicated that elevated ROS and a hyper polarized mitochondrion evinced in SLE CD8^+^ T cells can be modulated by an antioxidant. This is in line with other studies showing that NAC could plasticize cells towards an inflammatory or anti-inflammatory phenotype, depending upon the initial oxidative stress or disease condition affecting the redox system of the immune cells [24, 39, 42–46]. We also observed that there was no difference in the expression of anti-inflammatory cytokine, IL-10 either in menadione or NAC treated cells. Further, with PMA/Ion stimulation, IL-10 production showed minimal variation suggesting that this could be an inherent disease characteristic and could be attributed to the cells being highly inflammatory. The significantly higher levels of IFN-γ, IL-12p40, TNF-α, IL-6, IL-1β, IL-17A, IL-17E, IL-21and IL-15 could easily down regulate not only IL-10 response but also all of the Th2 cytokine responses too. Our study cohort mostly found naïve and memory cells in SLE patients while healthy controls had effector cells. We did see an increase in cytotoxic molecules such as Granzyme-B and Perforin-C in SLE patients but those molecules did not show any modulation with oxidant or antioxidant treatment (data not shown) indicating these molecules may be refractory to Redox activity and or may work through other pathways.

We followed up the above findings with studies on STAT’s to confirm and consolidate that the above findings were indeed pathway driven [49]. When we analyzed for the status of p-STAT3 and p-STAT4 in SLE CD8^+^ T cells we observed that p-STAT3 did not show any changes with menadione or NAC treatment, and that only p-STAT4 decreased with NAC treatment and is also in line with our previous observation as p-STAT4 can modulate IFN-γ expression [50]. Since p-mTOR plays an important role in the activation of T cells based on redox axis, we also wanted to check its status with Menadione and NAC treatment in SLE CD8^+^ T cells. We observed that NAC was able to decrease the expression of p-mTOR when compared with both menadione and PMA/Ion treated cells indicating that NAC can downregulate oxidative stress in CD8^+^ T cells [39, 49]. Overall, our data is in line with a clinical trial study which showed NAC when given as adjunct therapy in SLE, decreases inflammation and disease activity (Ref) and also restores redox axis in T cells by inducing expression of FOXP3 and reducing p-mTOR[41]. We did not observe any changes in FOXP3 expression (data not shown) due to the fact that our cells were isolated and treated *ex vivo* but in the above-mentioned study, NAC was given orally and for a certain period of time and consequently the effect could be due to the mode, dosage and timing of NAC [41].

Our experiments with SLE CD8^+^ T cells confirmed our hypothesis that these cells were indeed hyper cytotoxic, hyper polarized and hyper inflammatory. Although these cells could be modulated for their cytokines, TF’s and MAPK’s, it led us to ask the question if these were experimental artifacts or true phenotype of SLE CD8^+^ T cells. Thus, to confirm that terminally differentiated CD8^+^ T cells were truly plastic, we differentiated CD8^+^ T cells towards a Tc1 phenotype and examined their Redox sensitivity and consequential plasticity. Surprisingly we found that Menadione decreased the inflammatory cytokine expression such as IFN-γ and TNF-α and promoted the expression of anti-inflammatory cytokines in Tc1. Similarly, Eomes, a transcription factor responsible for maintaining the cytotoxic potential of CD8^+^ T cells also decreased with the addition of ROS while JAK-STAT proteins were also affected by oxidative stress. Further, we observed that the menadione increased the expression of p-STAT3 and reduced p-STAT4 expression. As p-STAT3 and p-STAT4 are known to regulate IL-10 and IFN-γ expression, we hypothesized that increased p-STAT3 could lead to higher IL-10 expression, whereas decrease in p-STAT4 could reduce IFN-γ expression. The expression of p-STAT3 and p-STAT4 reversed with the addition of ROS scavenger, NAC thus confirming that exogenous ROS is responsible for modulating p-STAT3/ p-STAT4 axis, thus governing inflammatory and anti-inflammatory phenotype of CD8^+^ T cells [48, 50, 51]. We observe that the trends with terminally differentiated Tc1 cells were diametrically opposite to the SLE CD8^+^ T cells and attribute the reversal to the disease process itself. As we evince significant levels of inflammatory cytokine in SLE patients and also know that the cytokine *milieu* can dictate activation and phenotype characteristics we furthered this line of thinking.

While the aberrant SLE CD8^+^ T cell had higher cytotoxic, inflammatory and ROS levels and was suggestive of an inflammatory cytokine *milieu* driven differentiation, this phenotype was not encountered in conventional Tc1 nor in normal controls. Interestingly, as the SLE plasma displayed higher IL-21 levels and with the known associated antibody pathology, we hypothesized that this cytokine from Tfh cells could be the potential modulator [5]. Thus, we terminally differentiated CD8^+^ T cells from healthy controls in presence of IL-21 along with IL-12 and IL-2, labeled as Tc21, and compared it to conventional Tc1. We observed that IL-21 did increase the cytotoxic and inflammatory potential of Tc21 cells when compared with Tc1 as was cellular ROS. However, they were not statistically significant indicating that although Tc21 had cytotoxic capabilities, but did not bear resemblance to the SLE CD8^+^ T cell. Thus, the cytokine milieu, disease stage, immune health etc. could play a role in transforming the CD8^+^ T cell [52, 53]. We previously reported from chronic hepatitis B patients a CD8 phenotype that was chronically suppressed, had different cytotoxic profile, inflammatory cytokines pattern, etc. than acute Hepatitis B patients and responded differently to NAC treatment displaying an increase in IFN-γ and decrease in IL10 and acute patients showing the reverse trend. It established the fact that cytokine and ROS in the microenvironment from the beginning of the pathology shapes the immune response of CD8^+^ T cells and certain steady phenotype of the disease could not be modulated by any treatments [17].

Although we understand that our study is based on a small sample size, but it essentially addresses the role played by the inflammatory and dysregulated immune environs in plasticizing CD8^+^ T cells. SLE CD8^+^ T cells bear characteristics that can only be partly mimicked in Tc21 cells concreting the above factors. But a study in a large cohort is required to understand which patients can be treated with NAC as an adjunct therapy as heterogeneity and endophenotypes exist in SLE patients. Additionally, our study essentially throws light into the crucial aspect of redox axis playing a significant role in determining or modulating the phenotype of SLE CD8^+^ T cells.

## Data Availability

All the relevant data are given in the manuscript/ supplementary materials.

## Ethics Statement

The studies involving human participants were reviewed and approved by the Institutional Human Ethics Committee, Institute of Life Sciences. The Institutional Ethics Committee (IEC)/ Institutional Review Board (IRB) reference number is 74/HEC/18. Written consent informed to participate in this study was provided by the participants.

## Data Availability Statement

All the relevant data are given in the manuscript/ supplementary materials.

## Author Contributions

Experimental Design and Conceptualization – SD, SS and SM Sample Collection, processing and clinical scoring - JRP and PKB Flow Cytometry Experiments-SS, SM, SKS and RJ Cytokine Multiplexing-SS, GB and GMJ Data analysis-SS, GB and SD

