## Supplemental Table 1 for "IL-21, inflammatory cytokines and hyper polarized CD8^+^ T cells are central players in Lupus immune pathology"

| **REAGENT** | **CATALOGUE NUMBER** |
| --- | --- |
| **MILLIPLEX MAP Human TH17 Magnetic Bead Panel - Immunology Multiplex Assay** | **Millipore –**  **HTH17MAG-14K** |
| **Dynabeads™ Untouched™ Human CD8 T Cells Kit** | **Invitrogen-**  **11348D** |
| **BD FACS ^TM^ FACS Lysing Solution** | **BD Biosciences-**  **349202** |
| **Dynabeads™ Human T-Activator CD3/CD28 for T Cell Expansion and Activation** | **Gibco-**  **11132D** |
| **Human TruStain FcX™ (Fc Receptor Blocking Solution)** | **Biolegend-**  **422302** |
| **BD Cytofix/Cytoperm™** | **BD Biosciences-**  **554714** |
| **eBioscience™ Foxp3 / Transcription Factor Staining Buffer Set** | **Invitrogen-**  **11348D** |
| **CellROX™ Deep Red Flow Cytometry Assay Kit** | **Invitrogen-**  **C10491** |
| **MitoTracker™ Red CMXRos Dye, for flow cytometry** | **Invitrogen-**  **M46752** |
| **Menadione** | **Sigma Aldrich**  **M9429** |
| ***N*-Acetyl-L-cysteine (NAC)** | **Sigma Aldrich**  **A9165** |
| **Anti-Human CD3 BV510** | **BD Biosciences-**  **740187** |
| **Anti-Human CD4 PECy7** | **BD Biosciences-**  **557852** |
| **Anti-Human CD8 AF700/BUV395** | **BD Biosciences-**  **561453/** **563795** |
| **Anti-Human CD278 (ICOS) PE** | **BD Biosciences-**  **557802** |
| **Anti-Human CXCR5(CD185) AF647** | **BD Biosciences-**  **558113** |
| **Zombie Fixable Viability™ Sampler Kit** | **Biolegend-**  **423117** |
| **Anti-Human pSTAT3 PE** | **BD Biosciences-**  **562072** |
| **Anti-Human pSTAT4 APC** | **Invitrogen-**  **17-9044-42** |
| **Anti-Human p-mTOR PECy7** | **Invitrogen-**  **25-9718-42** |
| **Anti-Human IFN-γ BV650/PECy7/AF-647** | **Biolegend/Invitrogen**  **502537/ 25-7319-41** |
| **Anti-Human IL-10 PE/BV 421** | **Invitrogen/Biolegend-**  **12-7108-82/** **501421** |
| **Anti-Human TNF-α BUV395/AF700** | **BD Biosciences-**  **563996/** **561023** |
| **Anti-Human Eomesdermin PECF594** | **BD Biosciences-**  **567167** |
| **Anti-Human Granzyme B PE** | **Biolegend-**  **372208** |
| **Anti-Human Perforin C AF594** | **Biolegend-**  **308124** |
| **Anti-Human T-bet PECy7** | **Invitrogen-**  **25-5825-82** |
| **Anti-Human Foxp3 PECy7/R718** | **Invitrogen/ BD Biosciences-**  **25-4777-42/** **566936** |
| **Anti-Human CD45 RO BV 605** | **BD Biosciences-**  **562791** |
| **Anti-Human CD45 RA BUV 496** | **BD Biosciences-**  **750258** |
| **FlowJO Version 10.8** | **BD Biosciences** |
| **Graph Pad Prism 9** | **Dotmatics Pvt Ltd.** |

**.**

**S Table 1:- Reagents and software used in the study**
