## Supplemental Figures for "IL-21, inflammatory cytokines and hyper polarized CD8^+^ T cells are central players in Lupus immune pathology"

### Slide 1
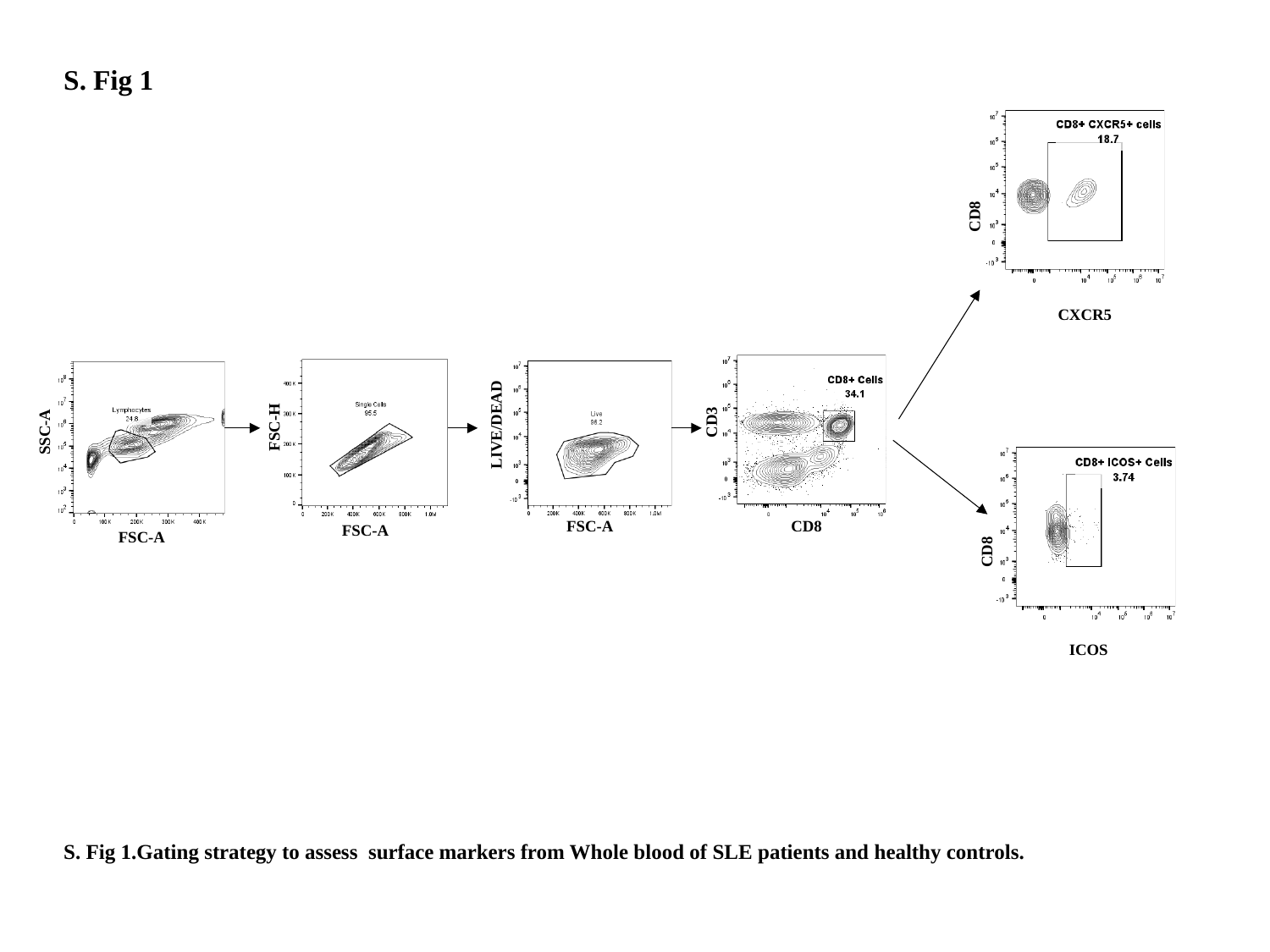

S. Fig 1
CD8
CXCR5
CD8
ICOS
CD3
FSC-H
LIVE/DEAD
SSC-A
FSC-A
CD8
FSC-A
FSC-A
S. Fig 1.Gating strategy to assess surface markers from Whole blood of SLE patients and healthy controls.

### Slide 2
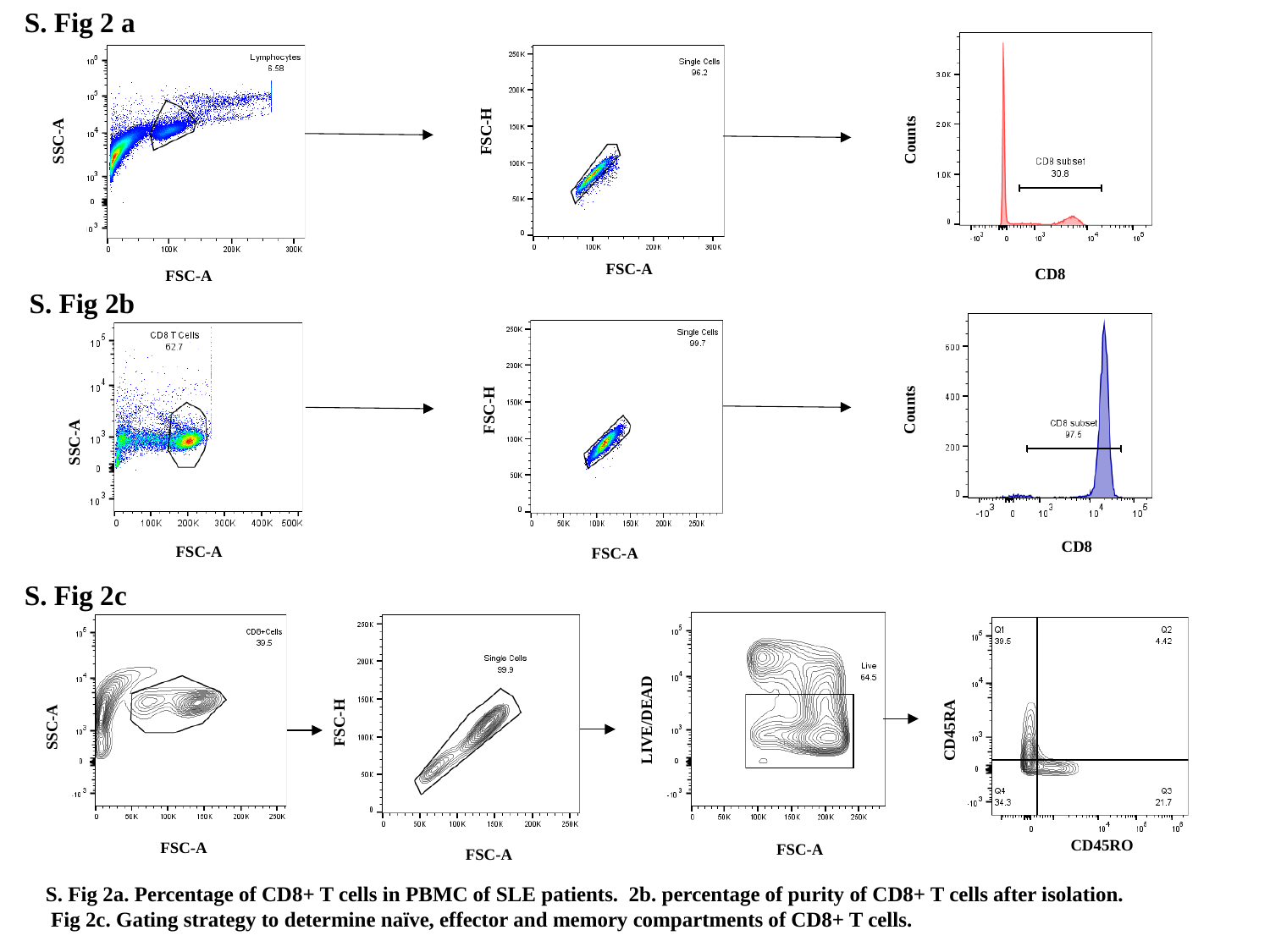

S. Fig 2 a
FSC-H
SSC-A
Counts
FSC-A
CD8
FSC-A
S. Fig 2b
FSC-H
Counts
SSC-A
CD8
FSC-A
FSC-A
S. Fig 2c
CD45RA
FSC-H
LIVE/DEAD
SSC-A
CD45RO
FSC-A
FSC-A
FSC-A
S. Fig 2a. Percentage of CD8+ T cells in PBMC of SLE patients. 2b. percentage of purity of CD8+ T cells after isolation.
 Fig 2c. Gating strategy to determine naïve, effector and memory compartments of CD8+ T cells.

### Slide 3
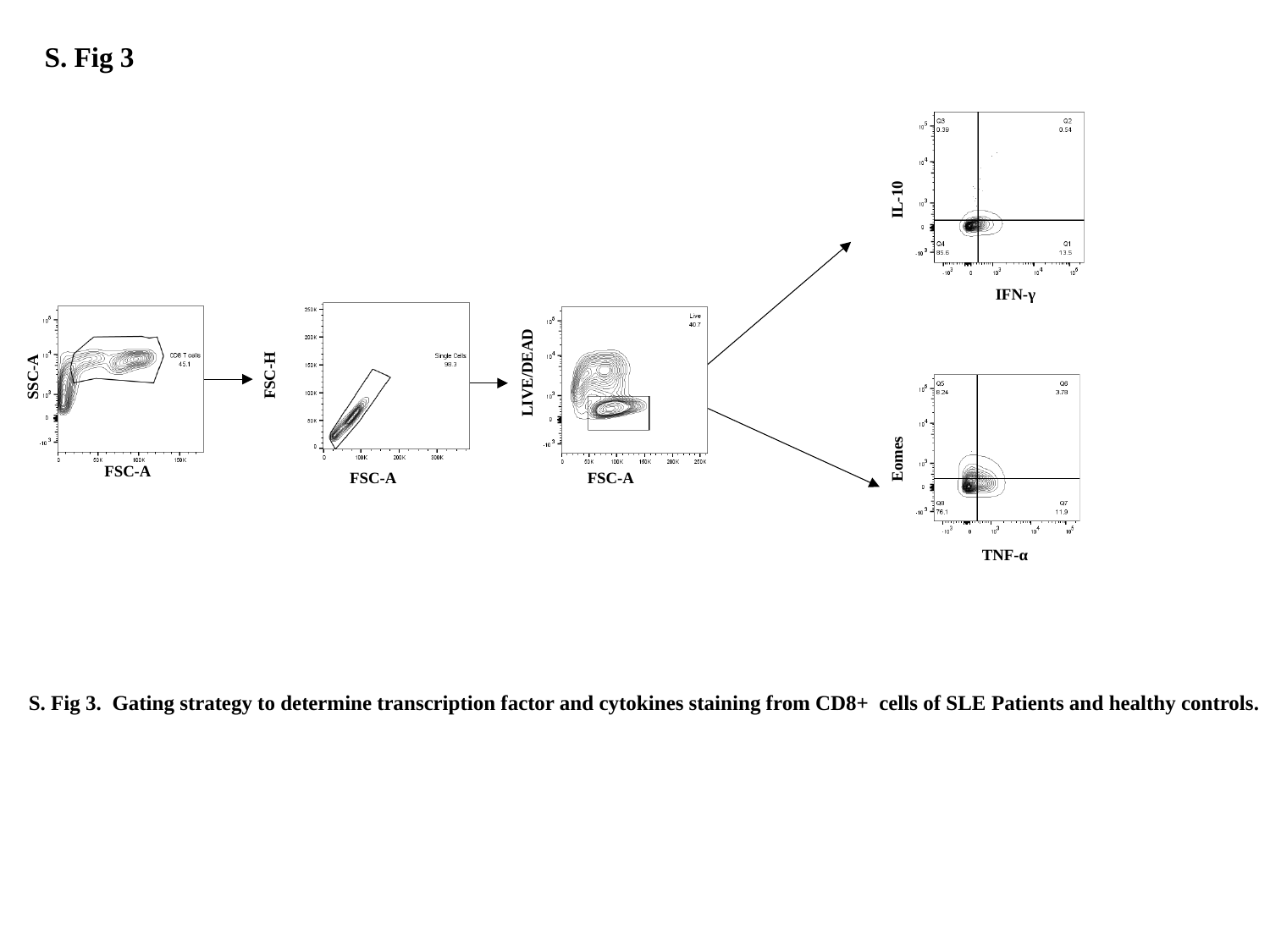

S. Fig 3
IL-10
IFN-γ
SSC-A
FSC-H
LIVE/DEAD
Eomes
FSC-A
FSC-A
FSC-A
TNF-α
S. Fig 3. Gating strategy to determine transcription factor and cytokines staining from CD8+ cells of SLE Patients and healthy controls.

### Slide 4
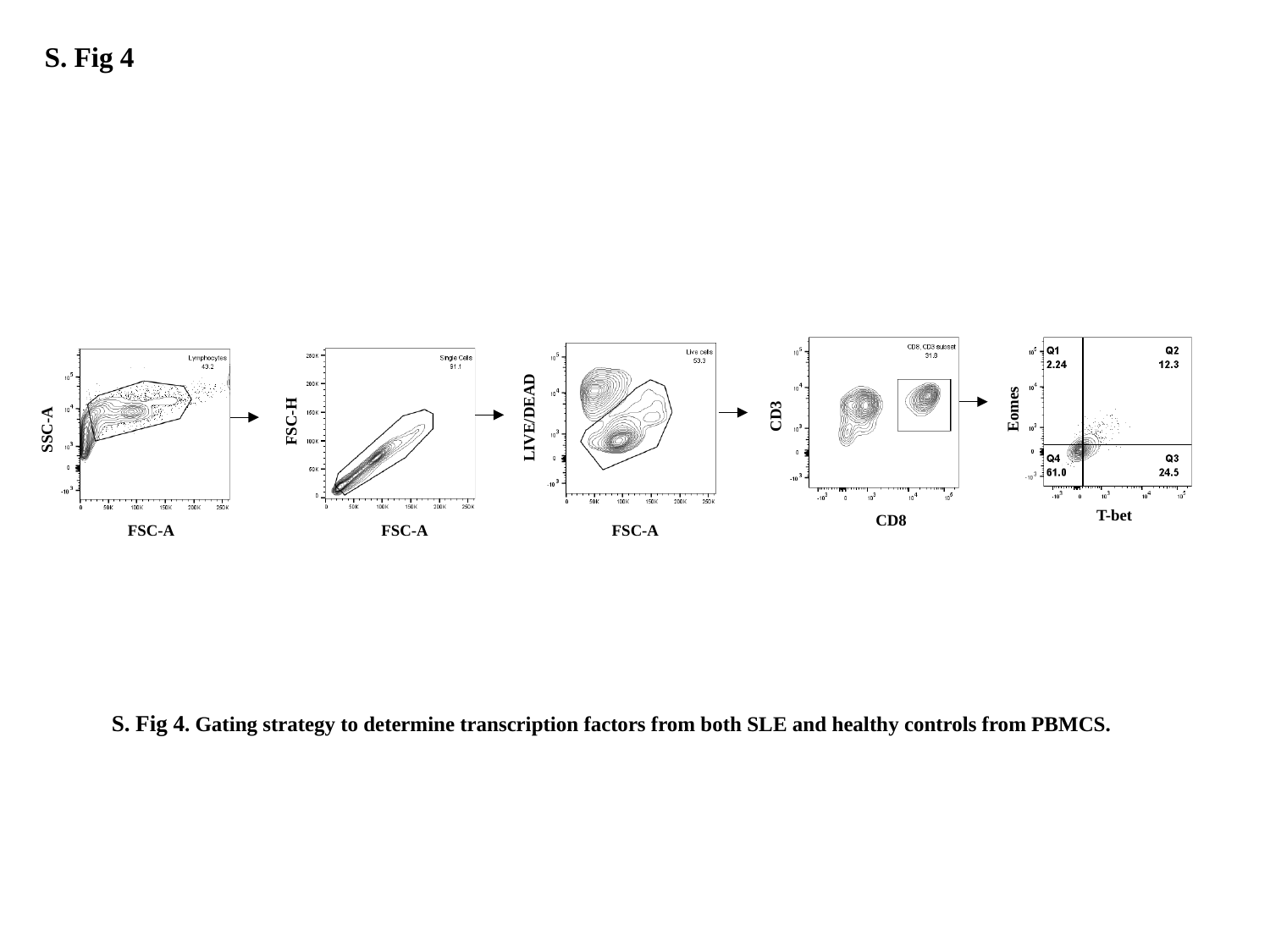

S. Fig 4
CD3
Eomes
LIVE/DEAD
FSC-H
SSC-A
T-bet
CD8
FSC-A
FSC-A
FSC-A
S. Fig 4. Gating strategy to determine transcription factors from both SLE and healthy controls from PBMCS.

### Slide 5
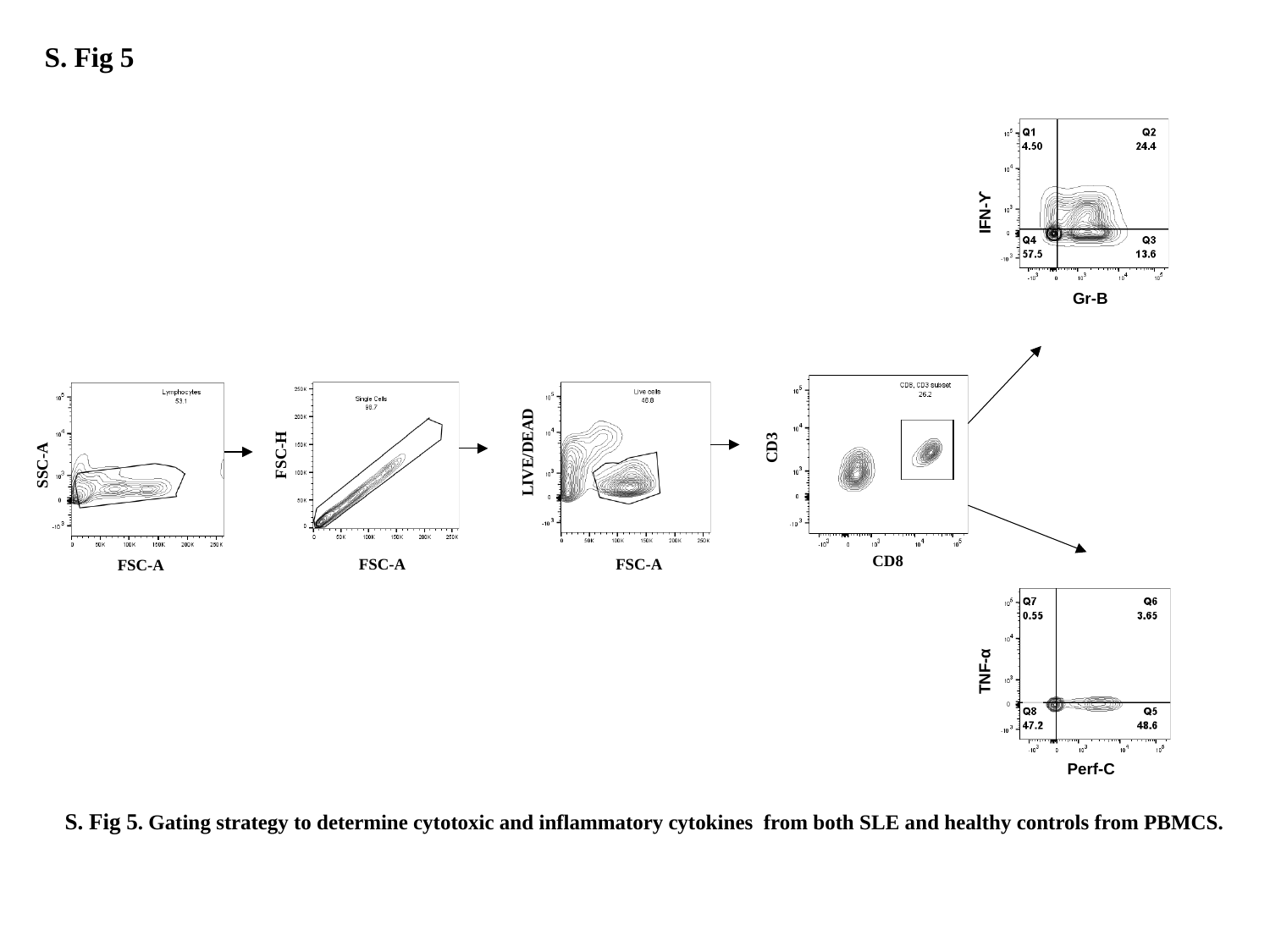

S. Fig 5
IFN-ϒ
Gr-B
CD3
FSC-H
LIVE/DEAD
SSC-A
CD8
FSC-A
FSC-A
FSC-A
TNF-α
Perf-C
S. Fig 5. Gating strategy to determine cytotoxic and inflammatory cytokines from both SLE and healthy controls from PBMCS.

### Slide 6
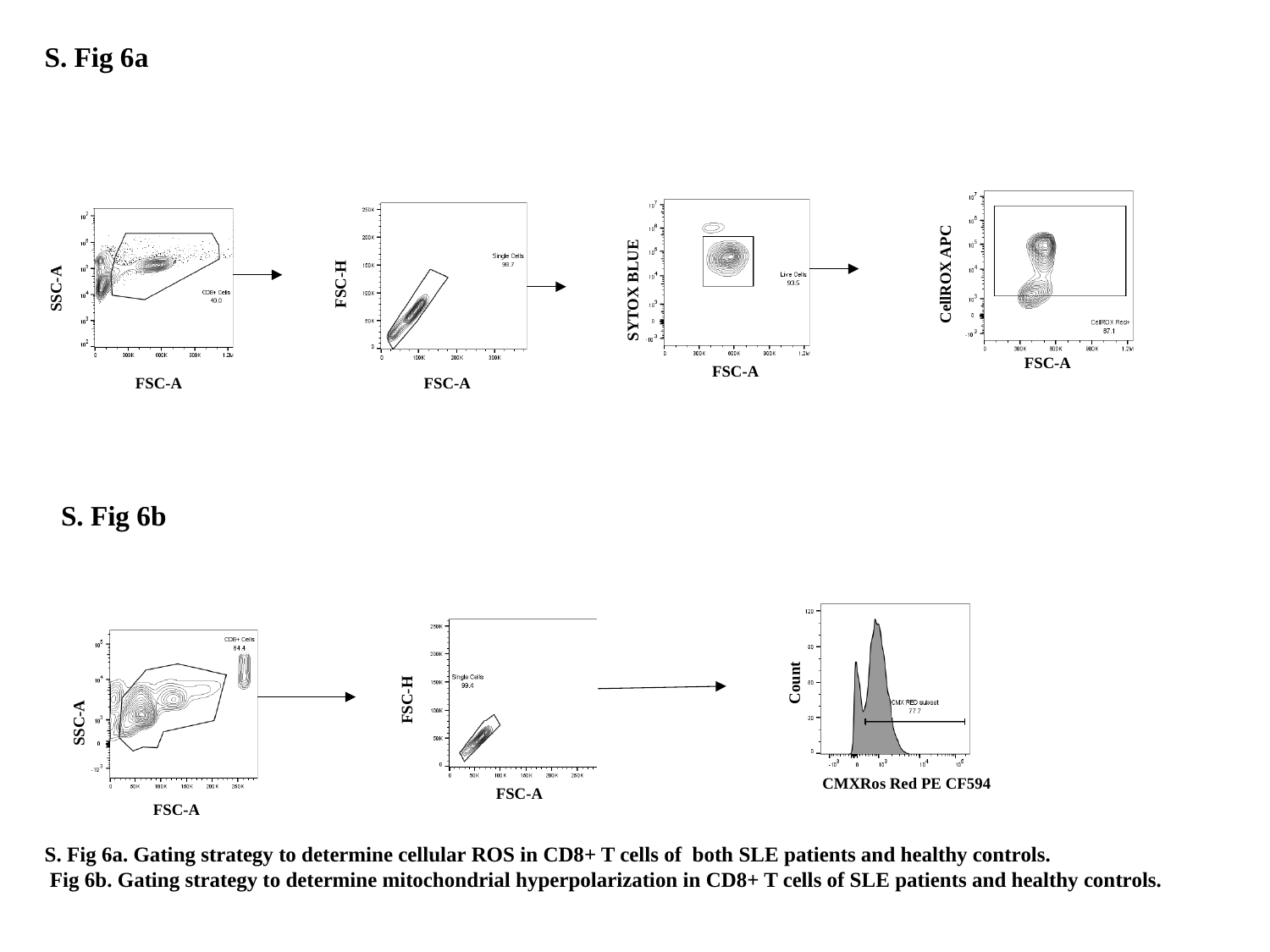

S. Fig 6a
FSC-H
FSC-A
SYTOX BLUE
FSC-A
SSC-A
FSC-A
CellROX APC
FSC-A
S. Fig 6b
Count
FSC-H
SSC-A
CMXRos Red PE CF594
FSC-A
FSC-A
S. Fig 6a. Gating strategy to determine cellular ROS in CD8+ T cells of both SLE patients and healthy controls.
 Fig 6b. Gating strategy to determine mitochondrial hyperpolarization in CD8+ T cells of SLE patients and healthy controls.

### Slide 7
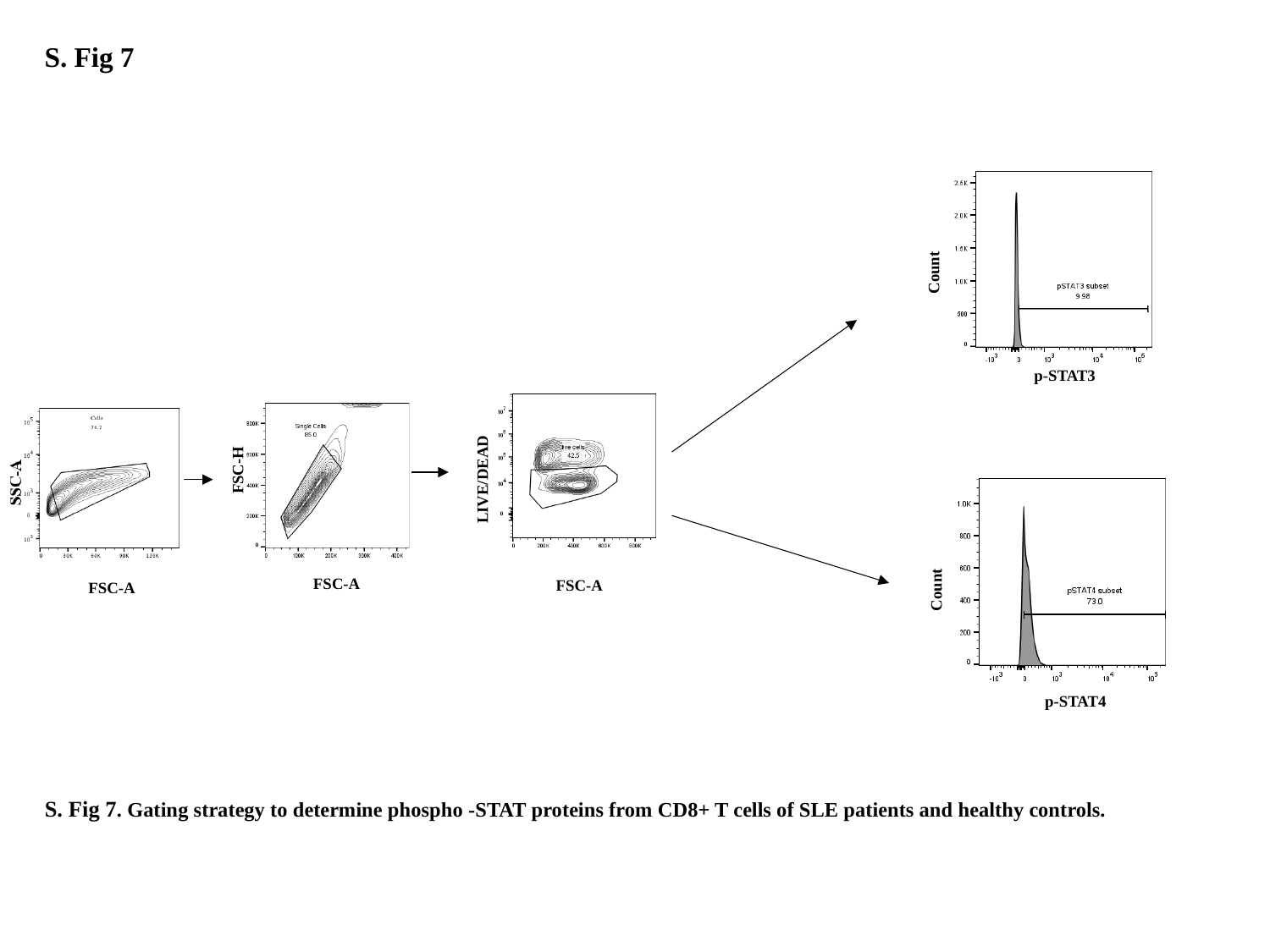

S. Fig 7
Count
p-STAT3
FSC-H
FSC-A
FSC-A
LIVE/DEAD
FSC-A
Count
p-STAT4
#### S. Fig 7. Gating strategy to determine phospho -STAT proteins from CD8+ T cells of SLE patients and healthy controls.

### Slide 8
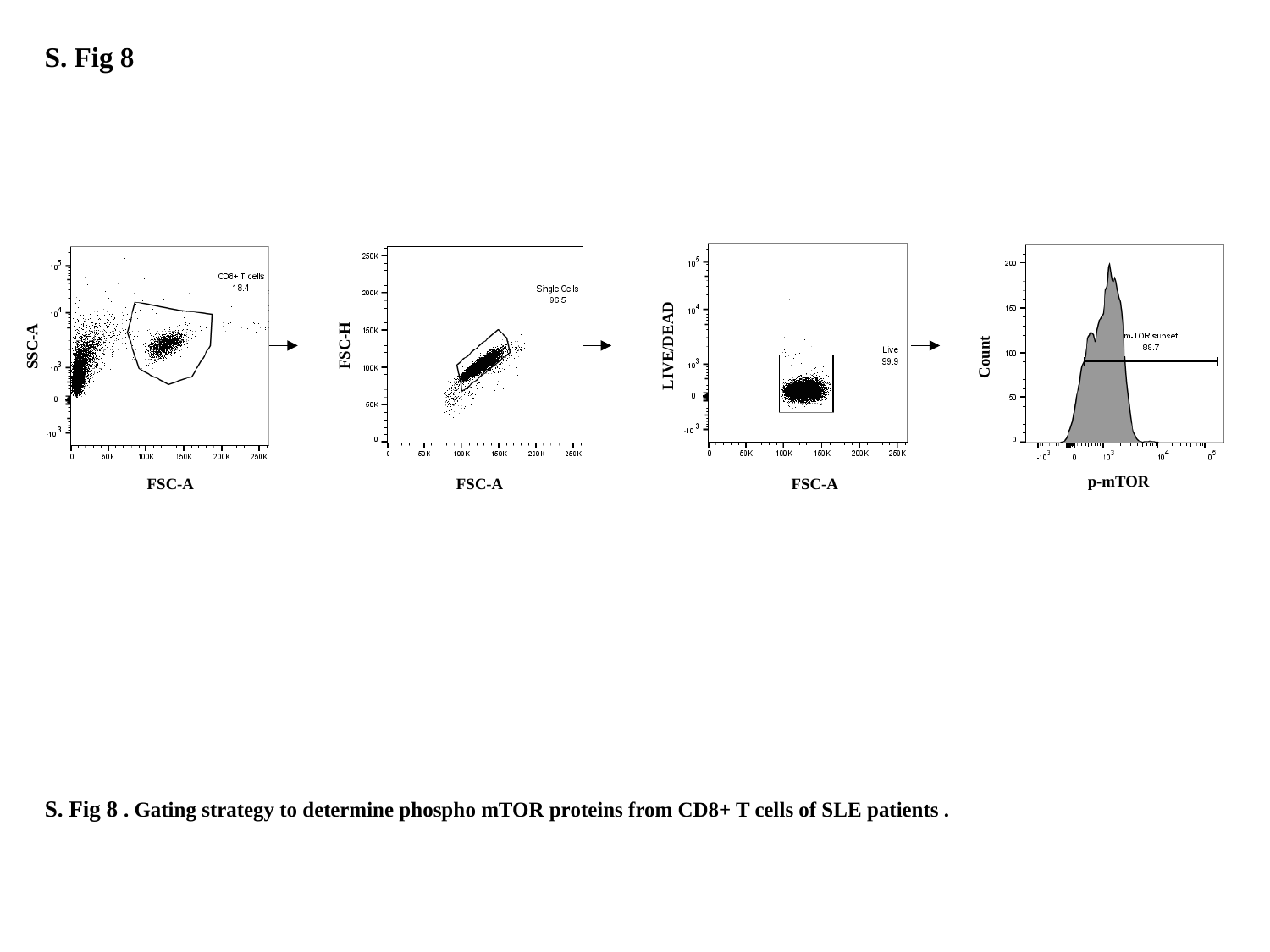

S. Fig 8
SSC-A
FSC-H
LIVE/DEAD
Count
p-mTOR
FSC-A
FSC-A
FSC-A
#### S. Fig 8 . Gating strategy to determine phospho mTOR proteins from CD8+ T cells of SLE patients .

### Slide 9
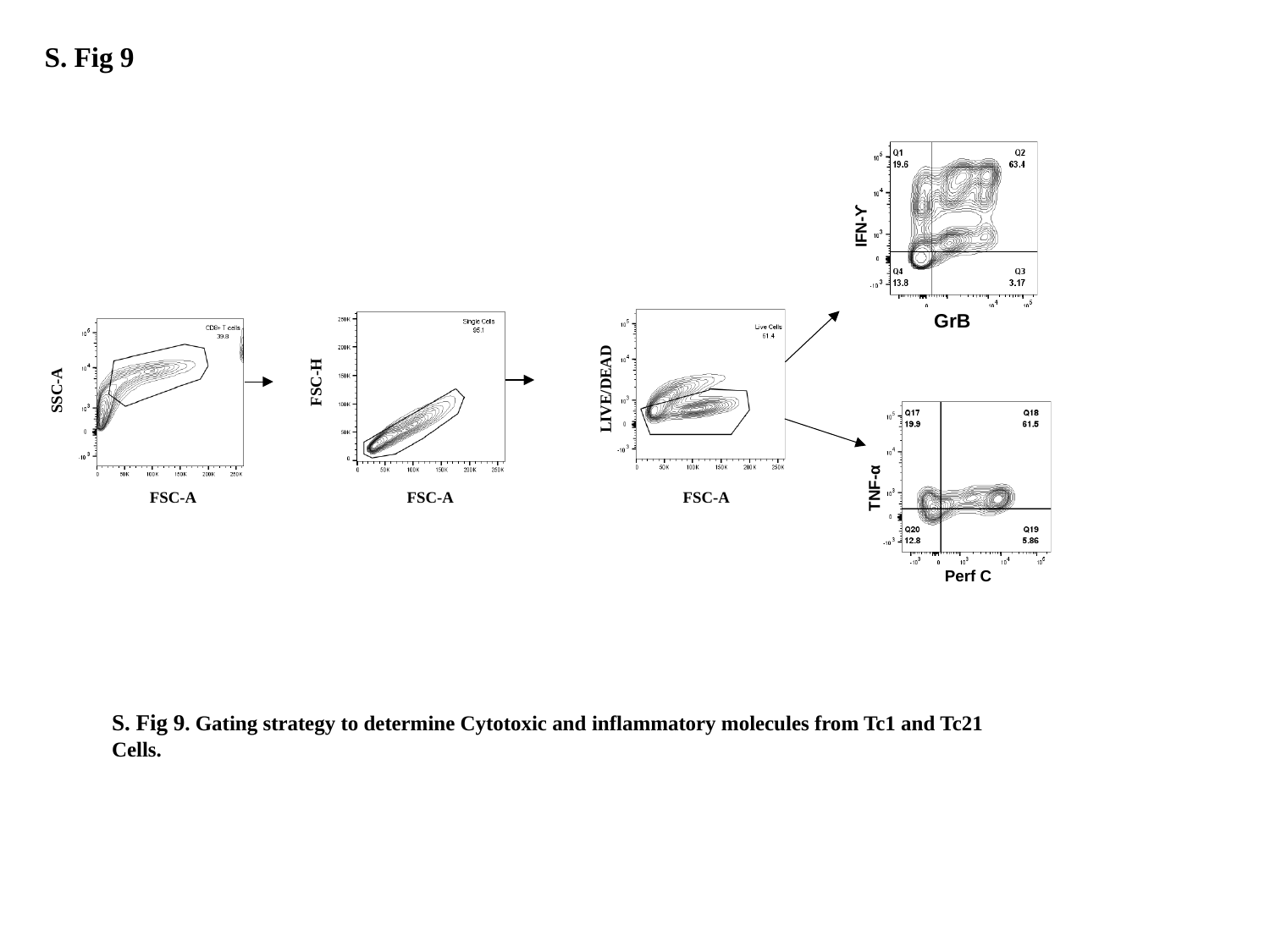

S. Fig 9
IFN-ϒ
FSC-H
SSC-A
LIVE/DEAD
FSC-A
FSC-A
FSC-A
GrB
TNF-α
Perf C
S. Fig 9. Gating strategy to determine Cytotoxic and inflammatory molecules from Tc1 and Tc21 Cells.
